# Time-Lapse Quantitative Analysis of Drying Patterns and Machine Learning for Classifying Abnormalities in Sessile Blood Droplets

**DOI:** 10.1101/2024.05.15.24307398

**Authors:** Anusuya Pal, Miho Yanagisawa, Amalesh Gope

## Abstract

When a colloidal droplet dries on a substrate, a unique pattern results from multi-facet phenomena such as Marangoni convection, capillary flow, mass transport, mechanical stress, colloid-colloid, and colloid-substrate interactions. Even under uniform conditions (surface wettability, humidity, and temperature), slight differences in the initial colloidal composition alter the drying pattern. This paper shows how the evolving patterns during drying in the sessile droplets depend on the initial composition and are crucial for assessing any abnormalities in the blood. To do so, texture statistics are derived from time-lapse images acquired during drying, and different traditional machine learning are applied. In addition, a neural network analysis is performed on both images and their texture statistics. As the drying phenomena are correlated with the varying composition, these methods exhibit excellent performance in distinguishing blood abnormalities with an F1 score of over 97%. This indicates that analysis of time-lapse images during drying and their texture statistics, rather than conventional analysis using images at the final dry state, are crucial for classification. Our results highlight the potential of droplet drying as a low-volume, accurate, and simple screening tool for detecting the type and stage of any disease in bio-fluid samples, such as blood, urine, and saliva.

## I. INTRODUCTION

When colloidal droplets on a solid substrate are evaporated (dried) under uniform conditions (droplet size, shape, composition, surface properties, temperature, and relative humidity), a unique pattern appears [1]. This uniqueness is due to multi-facet phenomena such as Marangoni convection, capillary flow, mass transport, mechanical stress, colloid-colloid, and colloid-substrate interactions within the droplet as it dries [2]. So far, diverse patterns have been reported not only for colloids such as from polymers [3-7], to liquid crystals [8, 9]; but also for fluids containing various biological colloids such as from DNA [10-12], collagen [13, 1], and microtubules [15], to proteins [16, 17], all the way to microbes such as bacteria [18, 19], algae [20], and nematode worms [21]. These previous reports show that the patterns of such droplets are extremely sensitive not only to how they are dried but also to the initial state of the fluids before drying.

During drying, the evolving patterns strongly depend on many factors such as spreading and wetting [22], substrate roughness [23], substrate temperatures [24], relative humidity [25], impact energy [26], and initial contact angles [27]. In recent years, attempts to use dried blood patterns to aid in crime scene investigation have attracted attention [28, 29]. It has also been suggested that the patterns in dried blood change depending on the state of the matrix contained in the blood, for example, blood plasma with [30] and without clotting factors [31]. Even at the laboratory level, variations in the patterns are observed when adding buffer [32] or deionized water [33], altering the composition of the blood. As the physical understanding of the drying patterns of bio-fluids, including blood, plasma, urine, saliva, etc., advances, it will be possible to estimate their states from the drying patterns, which is expected to contribute significantly to detecting various diseases [34 -35] and bio-medicine.

A challenge in achieving the above goals is the classification of dry droplets with complex compositions. In most cases, the shape, size, counts, composition, and morphology of the bio-fluids components vary, depending on the stage and the type of the disease. Until now, patterns have been analyzed using machine learning algorithms (ML) and neural networks (NN) [37-40]. The conventional analysis target is the images or just data in the form of contact angle, relative humidity, and temperature taken in the final drying stage. Therefore, some problems demand a substantial number of samples, and the diagnostic methods based on dried droplets, as discussed in [2, 41], are still in their nascent stages due to individual variations and their unsuitability for combining samples from multiple patients. As a result, there is an urgent need to identify more efficient strategies to translate this technology into clinical applications, thereby amplifying its potential to positively impact public health outcomes.

This paper presents an innovative analysis method by combining time-lapse images of evolving patterns during the drying process, quantitative image textural analysis, and data-driven approaches, such as traditional ML and NN. This approach holds the potential to unlock new insights and facilitate more accurate pattern recognition, offering promising avenues for advancements in data science. This method makes it possible to classify drying patterns with high accuracy even when the amount of fluids is less, compared to conventional analysis that uses only images of the final dried states. Although our analysis is based on an arbitrary classification of blood abnormalities in a bio-mimetic sense, the textural statistics can differentiate the biophysical changes within the blood components during the drying process. Therefore, this study is an innovative and valuable approach to identifying different bio-fluids of unknown compositions, with the hope of classifying diseased samples by correlating the drying patterns and varying fluids components. Furthermore, our approach to distinguishing between different types of blood cells or detecting abnormal cell morphology indicative of disease involves biotechnological techniques (microscopy and image analysis) with data science (time-lapse pattern recognition) to develop diagnostic tools. Figure 1 illustrates a schematic depicting the fusion of drying experiments and ML methodologies to classify different abnormalities in human blood.

**FIG. 1.**
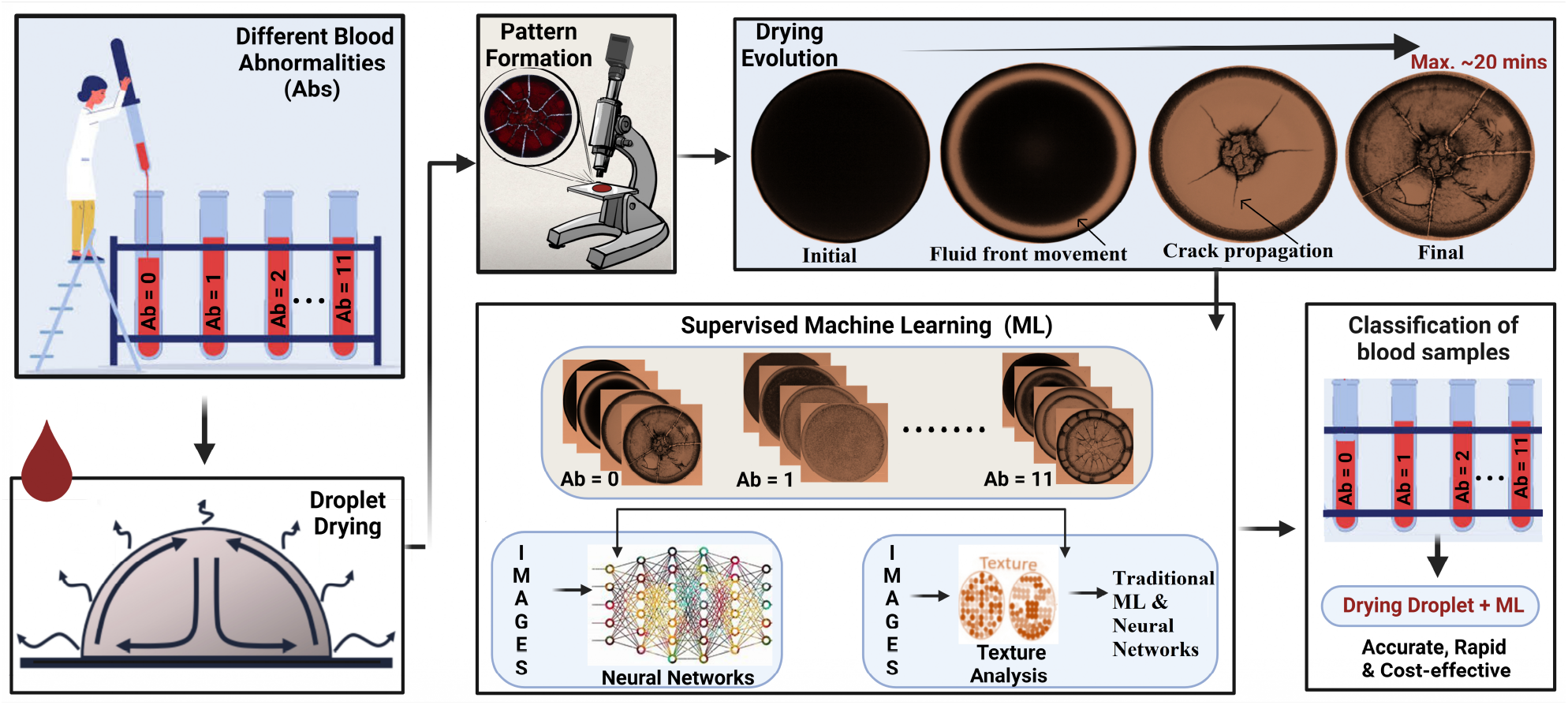
Illustration of innovative analysis to classify different abnormalities in human blood by combining time-lapse images of evolving patterns during the drying process, quantitative image textural analysis, and data-driven approaches, such as traditional machine learning (ML) and Neural Networks (NN). Our study breaks new ground by utilizing the evolving dynamics of pattern formation during drying as a unique fingerprint, moving beyond focusing solely on dried morphologies. The combination of drying pattern dynamics and ML helps us find a low-volume, rapid, accurate, and cost-effective screening tool for diverse samples.

## II. EXPERIMENTAL METHODS

### A. Samples

The human blood (Catalog number 7203706) was acquired from Lampire Biological Laboratories, USA, and contained Na-Citrate anticoagulant. The company strictly followed the informed written consent from all participants and the necessary FDP rules and regulations. Therefore, approval from a national or institutional ethics board and the committee was not required before this multi-disciplinary research. The study employed 1x phosphate buffer saline (PBS, BP243820, Fisher BioReagents, USA) and de-ionized water (Millipore, with a conductivity of 18.2 MΩ.cm).

### B. Sample preparations

To modify the concentration of whole blood (initially at 0% by volume), we employed a dilution strategy involving the addition of 1x PBS and de-ionized water. Notably, 1x PBS contained a composition of 0.137 M NaCl, 0.0027 M KCl, and 0.119 M phosphates while maintaining a consistent pH within the range of 7.3 − 7.5. Diverse blood samples were prepared to cover a spectrum of concentrations, ranging from 12.5 to 75% (by volume). Importantly, all experimental procedures were executed soon after sample preparation. This rapid processing ensured that lysis of various blood components did not transpire before the initiation of the drying process. A total of eleven blood samples was prepared, encompassing various compositions: healthy (0 (v/v) %), blood mixed with PBS (12.5, 25, 50, 62, and 75 (v/v) %), and blood mixed with water (12.5, 25, 50, 62, and 75 (v/v) %). Each of these samples, with a volume of approximately ∼1*μ*L, was carefully pipetted onto a microscopic coverslip (Catalog number 48366 -045, VVR, USA). This action resulted in circular droplets, each droplet radius of ∼1 mm. All experimental procedures were carried out under standard ambient conditions, characterized by a room temperature of ∼25°C, and an RH of ∼50%. To ascertain the reproducibility of our findings, each experiment was conducted in triplicate.

### C. Image acquisition

The drying process of blood droplets was observed using bright field microscopy (Amscope, USA) with a 5× objective lens. A digital camera (8-bit, MU300, Amscope, USA) was connected to the microscope for time-lapse image capture at two-second intervals after droplet deposition. Images were captured at a resolution of 3664 × 2748 pixels. The lamp intensity remained constant throughout the drying process to minimize background fluctuations. A calibration slide was used to convert pixels to real-space length scales. All images were transformed into 8-bit grayscale images for improved visualization. Typically, the time-lapse sequence, spanning two seconds per interval, yielded 400-600 images capturing the entire drying process for a droplet with a radius of ∼1 mm, the volume of ∼1*μ*L, dried under ambient conditions of T ∼25°C and RH ∼50%.

### D. Image processing

The ImageJ software [42] was utilized to select a circular region of interest (ROI) using the *oval tool*. Gray values in the 8-bit images ranged from 0 to 255. Textural features, including first-order statistics (FOS) and gray-level co-occurrence matrix (GLCM) attributes, were extracted. FOS parameters encompassed mean, standard deviation, kurtosis, and skewness of the droplet. GLCM parameters encompassed angular second moment, contrast, correlation, inverse difference moment, and entropy. GLCM parameters were computed using the *Texture Analyzer plugin* in ImageJ [1]. Mathematical definitions for each feature are detailed in the supplementary section. Nine textural parameters were used as features in traditional ML, and artificial neural network approaches for classifying distinct blood samples.

### E. Machine learning (ML)

Approximately 550 images were captured throughout the drying process for each blood sample. The total number of images (dataset) ∼ 6 × 10^3^. Consequently, we generated an equivalent number of datasets, as we extracted FOS and GLCM textural features from each image. This dataset can be denoted as *D* = (*x*_1_, *y*_1_), (*x*_2_, *y*_2_), … (*x*_*n*_, *y*_*n*_), with *n* representing the total dataset size (approximately 0). Each *x* comprises a feature vector consisting of nine elements: mean, standard deviation, kurtosis, skewness, angular second moment, contrast, correlation, inverse difference moment, and entropy. Correspondingly, *y* designates the corresponding class, encompassing 11 distinct types of blood samples. All ML implementations within this study are supervised learning. The dataset (*D*) was divided randomly into two mutually exclusive groups: *D*_*train*_ and *D*_*test*_. Different ML algorithms were assessed using *D*_*train*_ and subsequently tested for performance on *D*_*test*_. Data manipulation was executed using Python s Pandas (version 0.24.2) and Numpy (version 1.16.4). The dataset was split into *D*_*train*_ and *D*_*test*_ using the *train*_*test*_*split*() function from the library [43]. A *test*_*size* of 0.3 was employed, indicating that 70% of *D* was designated as the training set, while the remaining 30% served as the testing set.

#### 1. Tradidiora ML

Five distinct traditional ML algorithms were employed in this study: Decision Tree (DT), Random Forest (RF), Support Vector Machine (SVM), K-Nearest Neighbors (KNN), and Naive Bayes (NB). Implementing these algorithms was facilitated through the *Scikit-Learn* library [43], which offers a Python interface for traditional ML methods. Prior to applying any ML technique, the data is scaled. Random Forest (RF) is built upon Decision Trees (DT). Each individual tree in the ensemble predicts outcomes, and a majority vote determines the final class prediction. RF employs bagging and feature randomness during tree construction, resulting in an uncorrelated forest of multiple trees. The RF classifier was instantiated using *RandomForestClassifier* from the *sklearn*.*ensemble* module. In our implementation, *n*_*estimators* was set to 400. Tree node splitting was determined based on *Shannon entropy* criteria. The RF architecture is depicted in Figure S1 of the supplementary section. Decision Trees (DT) follow a branching approach to illustrate possible decision outcomes. We utilized the *DecisionTreeClassifier* with *criterion = ‘entropy’*. This classifier was imported from *sklearn*.*tree*. K-Nearest Neighbors (KNN) uses a distance metric to classify data points. The *KNeighborsClassifier* was employed with parameters such as *n_neighbors* set to 5, *weights* as ‘distance’, and *metric* as ‘minkowski. These settings were imported from *sklearn*.*neighbors*. Support Vector Machine (SVM) is based on identifying a hyperplane and support vectors closest to the hyperplane to separate different classes. We used the *SVC ()* function from *sklearn*.*svm* for the analysis. Na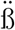ve Bayes (NB) leverages Bayes’ Probability Theorem to compute the probability of data belonging to a specific class. We imported *GaussianNB()* from *sklearn*.*naive_bayes* for classification purposes.

#### 2. Neural Networks (NN)

This study employed two types of neural networks: Artificial Neural Networks (ANN) and Convolutional Neural Networks (CNN). The primary distinction between these two lies in the data input format. While the data was directly utilized as input for ANN, the images were fed into the CNN architecture directly. To implement both ANN and CNN, we utilized *Keras* in conjunction with the *TensorFlow* library [44], Neural networks, whether ANN or CNN, operate on the concept of artificial neurons, which loosely mimic the neurons in the human brain. These networks establish probabilistic-weighted connections between inputs and output classes. The architecture of these networks is constructed and manipulated to predict output classes based on the inputs. Throughout the training process, the network s weighted connections are adjusted, aiming to achieve increasingly accurate output predictions. Training concludes after a sufficient number of iterations. Both ANN and CNN were developed using the *Sequential()* model from *Keras*. This involved the incorporation of layers such as *Dense, ReLU Activation*,, and *Dropout*. The output layer consisted of a *Dense layer* and a *Softmax Activation* layer. The model was compiled using parameters including *optimizer = adam’, loss = categorical_crossentropy’*, and *metrics = ‘accuracy’*. Detailed information regarding the architectures of both ANN and CNN can be found in the supplementary section.

#### 3. Evaluation of ML

Various metrics were employed to assess the efficacy of ML models in classifying blood samples. These metrics encompass: (i) 10-fold Cross-Validation: A 10-fold cross-validation procedure was applied, involving dataset shuffiing and division into ten groups. Each group was utilized as a test set, while the remaining nine served as the training set. The ML model was fitted on the training data, evaluated the test data, and derived an evaluation score. This process was repeated for all ten groups, enabling a comprehensive comparison. The *KFold* function from *sklearn*.*model_selection* was employed for this purpose. (ii) Confusion Matrix: The confusion matrix provides a matrix that contrasts actual and predicted values. This *m* × *m* matrix, where *m* denotes the number of classes (in this case, 11), showcases the relationship between true and predicted class labels. Diagonal elements correspond to correct predictions, while off-diagonal elements denote misclassifications. The *confusion_matrix* function from *sklearn*.*metrics* was employed. Typically, these matrix values are normalized and presented as a color map within the range of 0 to 1. (iii) Accuracy, Precision, Recall, and F1-Score: A series of classification metrics were computed, where accuracy represents the proportion of correctly predicted instances out of the total occurrences. Precision, Recall, and F1-score were also calculated. Mathematically, 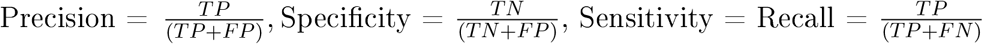, and 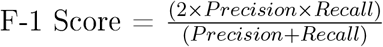. Here, True Positives (TP) indicate correctly predicted positive instances, False Positives (FP) represent incorrect positive predictions, True Negatives (TN) signify accurate negative predictions, and False Negatives (FN) correspond to inaccurate negative predictions. The *accuracy_score* and *classiffication_report* functions from *sklearn*.*metrics* were utilized to compute these metrics. Of note, the F1-score, which balances precision and recall, was employed for comparing ML performance.

## III. RESULTS AND DISCUSSIONS

### A. Visual inspection and textural analysis of the blood drying droplets

#### 1. Physics of evolving patterns in blood with and without abnormalities

The displayed images in Figure 3(I-II) are cropped from the original ones for improved visualization. We present time-lapse images depicting the drying process of a blood droplet (I) with no abnormalities and (II) with abnormalities (+ PBS) at a concentration of 12.5% (v/v). During the initial stages of the drying process, the behavior of the blood+PBS droplet closely resembles that of healthy blood. The first image was captured within 50-60 seconds after the droplet was deposited onto the coverslip (substrate). Initially, the droplet displays a uniformly dark texture with no noticeable changes. As time advances, a fluids front gradually advances from the periphery toward the center. The white dashed arc line illustrates this uniform front movement [see the blue panels of Figure 3(I-II)]. As the front progresses, a smooth gray texture appears behind it, forming a dark peripheral band (highlighted with orange arrows). Concurrently, the dark texture in the central region (indicated by a white dashed circular line) begins to lighten. Once the front vanishes, it becomes evident that two distinct regions exist-one is the central, and the other is the ring region.

As the drying progresses, differences between the two droplets are introduced, especially in the middle stage (depicted in green). Notably, at 12.5%, a dendritic structure begins to develop in the center of the droplet [see the red ovals in Figure 3(II)]. This growth distinguishes the drying process from that of healthy blood. Crack propagation initiates from the transition region of the central and ring regions, with white and yellow arrows indicating the direction of the cracks and symmetric stress fields, respectively. This is not found in healthy blood. In terms of the cracks, The white arrows sketch the propagation of the radial cracks, while *σ*_*l*_ and *σ*_*r*_ demonstrate the symmetric stress fields of these cracks (depicted with yellow arrows).

In contrast to the cracks in the central region of the healthy blood droplet [see Figure 3(I)], the cracks propagate toward the periphery. Unlike drying a healthy blood droplet, some cracks in blood+PBS propagate towards the central region, while others extend towards the periphery. Over time, cracks propagating towards the central region merge with the dendritic structure. In addition, the texture of the crack domains gets darkened only in the healthy blood [see the green dotted oval lines in Figure 3(I)].

Similar to the healthy blood droplet, we observe the branching and widening of radial cracks in the blood+PBS at 12.5% [see the yellow dashed rectangles in Figure 3(I-II)] as the final stage of the drying process. These visual observations provide valuable insights into the similarities and differences in the events occurring during the drying process in blood droplets with and without abnormalities. This suggests that the middle stage is the most salient stage of the unique pattern formation in such droplets.

#### 2. Texture analysis at different concentrations of abnormalities

Figure 2(I-II) showcases sequential image snapshots of blood drying droplets, depicting their initial, middle, and final stages when different volumes of (I) water and (II) PBS are added at the initial concentrations ranging from 12.5% to 75% (v/v). This sums up to eleven different types of abnormalities in the blood. Each image is timestamped relative to the total drying duration (*t*_*f*_). Here, we aim to comprehend the overall dynamics of blood samples, focusing on spatial and temporal behaviors critical for pattern emergence.

**FIG. 2.**
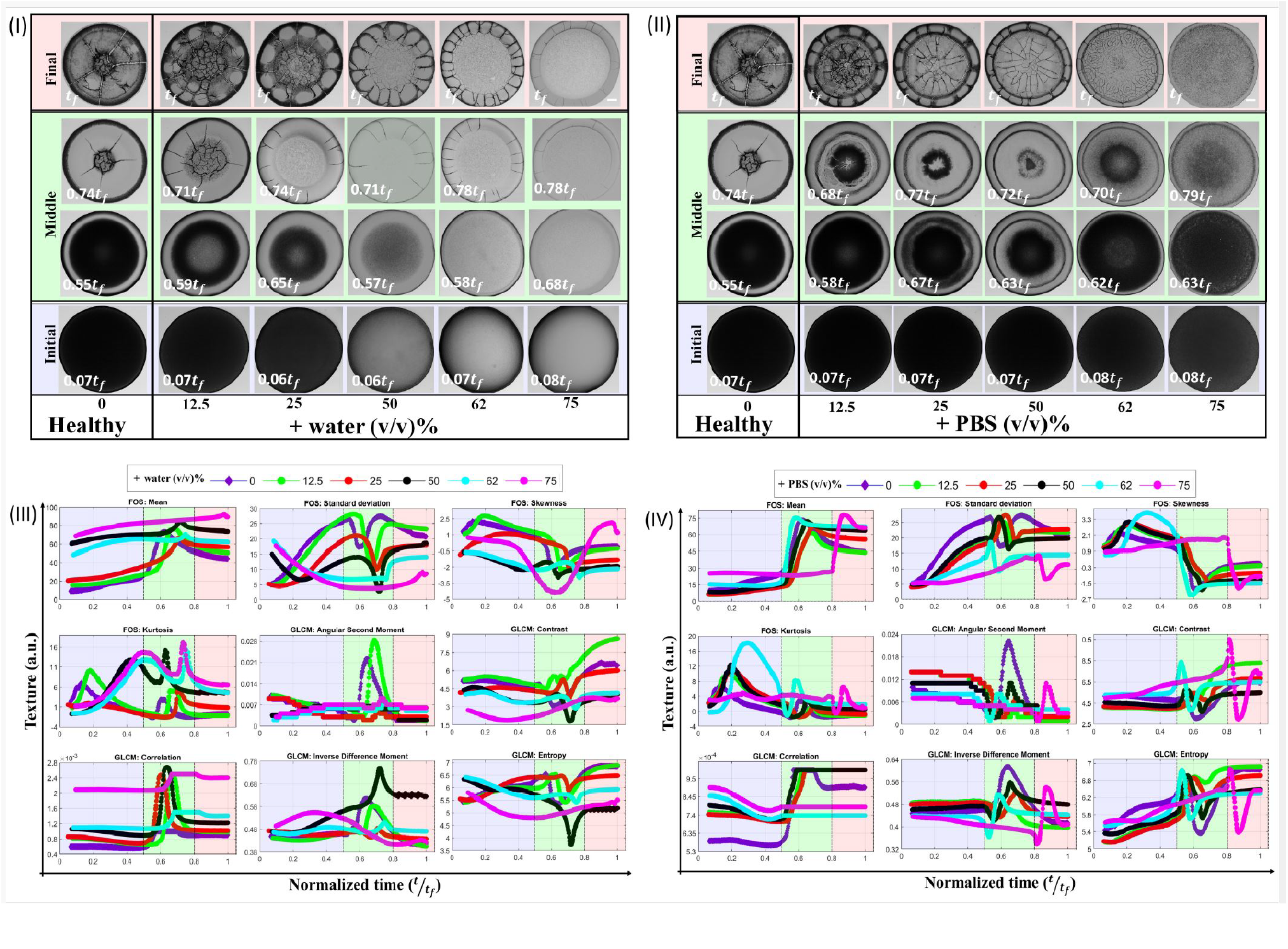
Time-lapse images capturing the drying process of blood droplets containing different abnormalities by varying volumes of added (I) Water and (II) Phosphate Buffered Saline (PBS), ranging from 12.5 to 75%(v/v). The 0%(v/v) corresponds to the drying of a healthy blood droplet, m. The final drying time, denoted as *t*_*f*_, signifies the point when no further visible changes occur in these droplets. The timestamp on each image is relative to *t*_*f*_. The scale bar of 0.2 mm is indicated in white color at the conclusion of the 75%(v/v) stage. Textural statistics comprising first-order statistics (FOS) and gray-level co-occurrence matrix (GLCM) are assessed from images captured at different concentrations, ranging from 12.5 to 75%(v/v). This assessment is plotted as a function of the normalized time 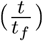 for (III) Added water and (IV) Added PBS. The FOS encompasses mean, standard deviation, skewness, and kurtosis, while the GLCM entails angular second moment, contrast, correlation, inverse difference moment, and entropy. Distinct stages of the drying process-initial, middle, and final-are represented by blue, green, and red colors, respectively.

**FIG. 3.**
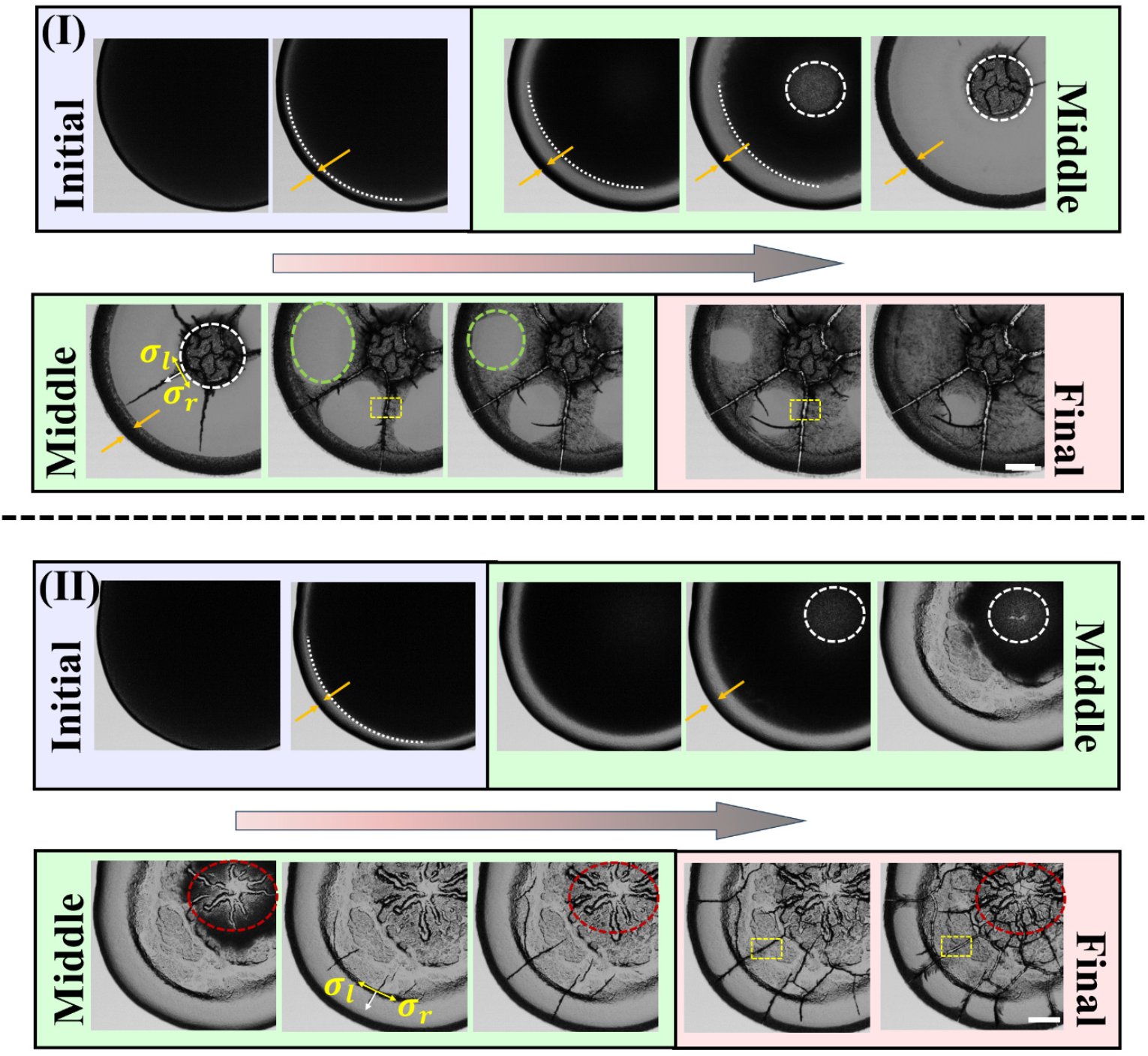
Time-lapse images of the blood droplets during the drying process (I) without any abnormalities and (II) with abnormalities (+ PBS) at 12.5% (v/v). The blue panel shows the deposited droplets, the formation of the peripheral band (depicted with orange arrows), and the movement of the fluid front. The green panel exhibits that the front moves uniformly from the periphery of the droplet (white dashed arc lines). The texture in the central region becomes lighter, outlined with a white dashed circle. The white arrows sketch the propagation of the radial cracks, while *σ*_*l*_ and *σ*_*r*_ demonstrate the symmetric stress fields of these cracks (depicted with yellow arrows). This panel emphasizes the differences between healthy and non-healthy blood. The green dotted oval line shows the texture change in the domains created by these radial cracks in the healthy blood. The red ovals mark the appearance of the dendrite structure in the central region of blood+PBS. The yellow dashed rectangles portray the widening of these radial cracks as the final stage of the drying process. The scale bar corresponds to 0.2 *mm*. Distinct stages of the drying process-initial, middle, and final-are represented by blue, green, and red colors, respectively. The gray arrows in (I-II) display the drying progression.

**FIG. 4.**
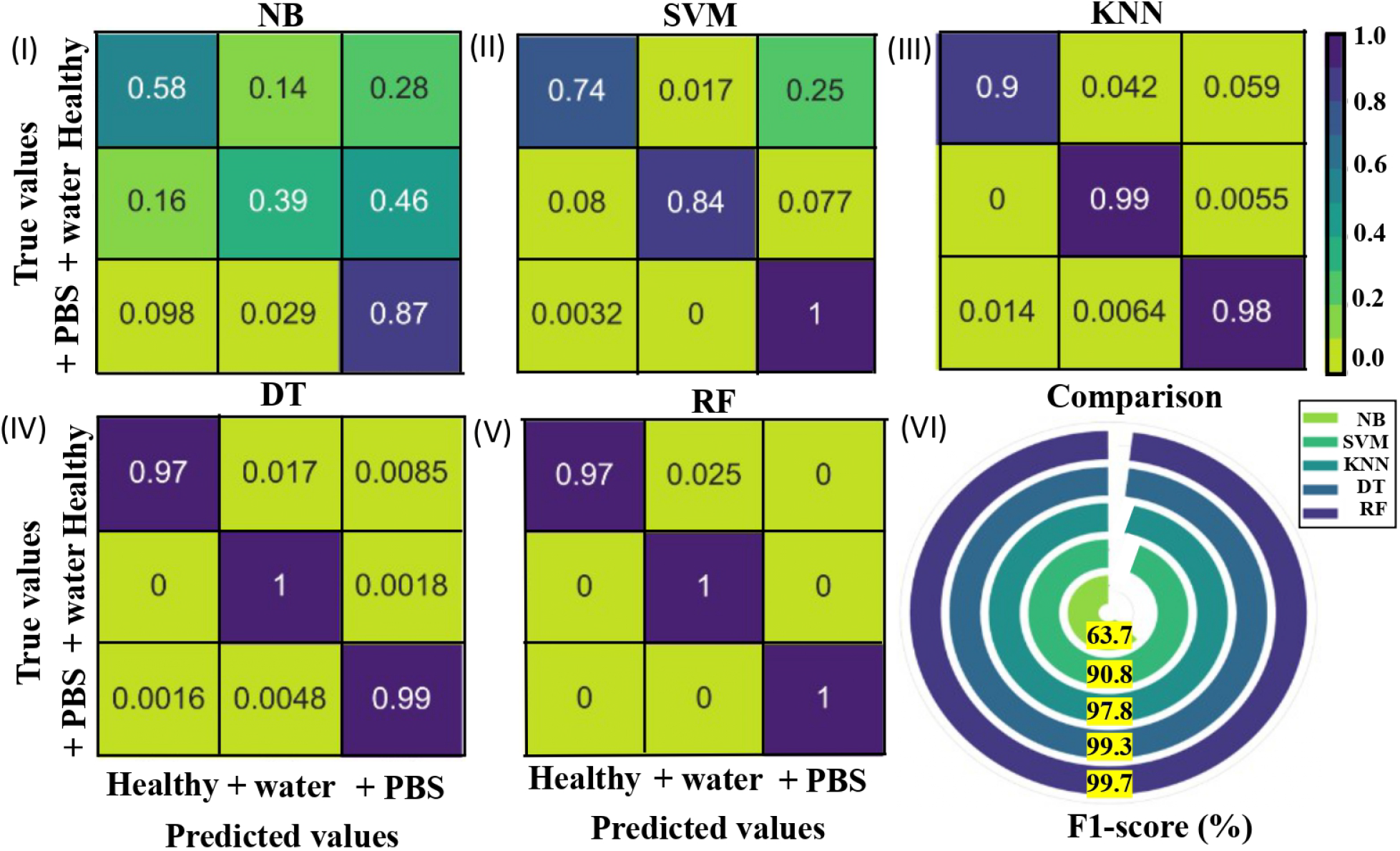
Normalized confusion matrix illustrating the performance of various ML: (I) Na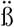ve Bayes (NB), (II) Support Vector Machine (SVM), (III) K-Nearest Neighbors (KNN), (IV) Decision Tree (DT), and (V) Random Forest (RF). These MLs are employed to classify healthy blood droplets and blood droplets with added water or phosphate buffer saline (PBS). (VI) The radial plot depicting the average F1-score in percentage provides a comparative assessment of each ML’s performance.

**FIG. 5.**
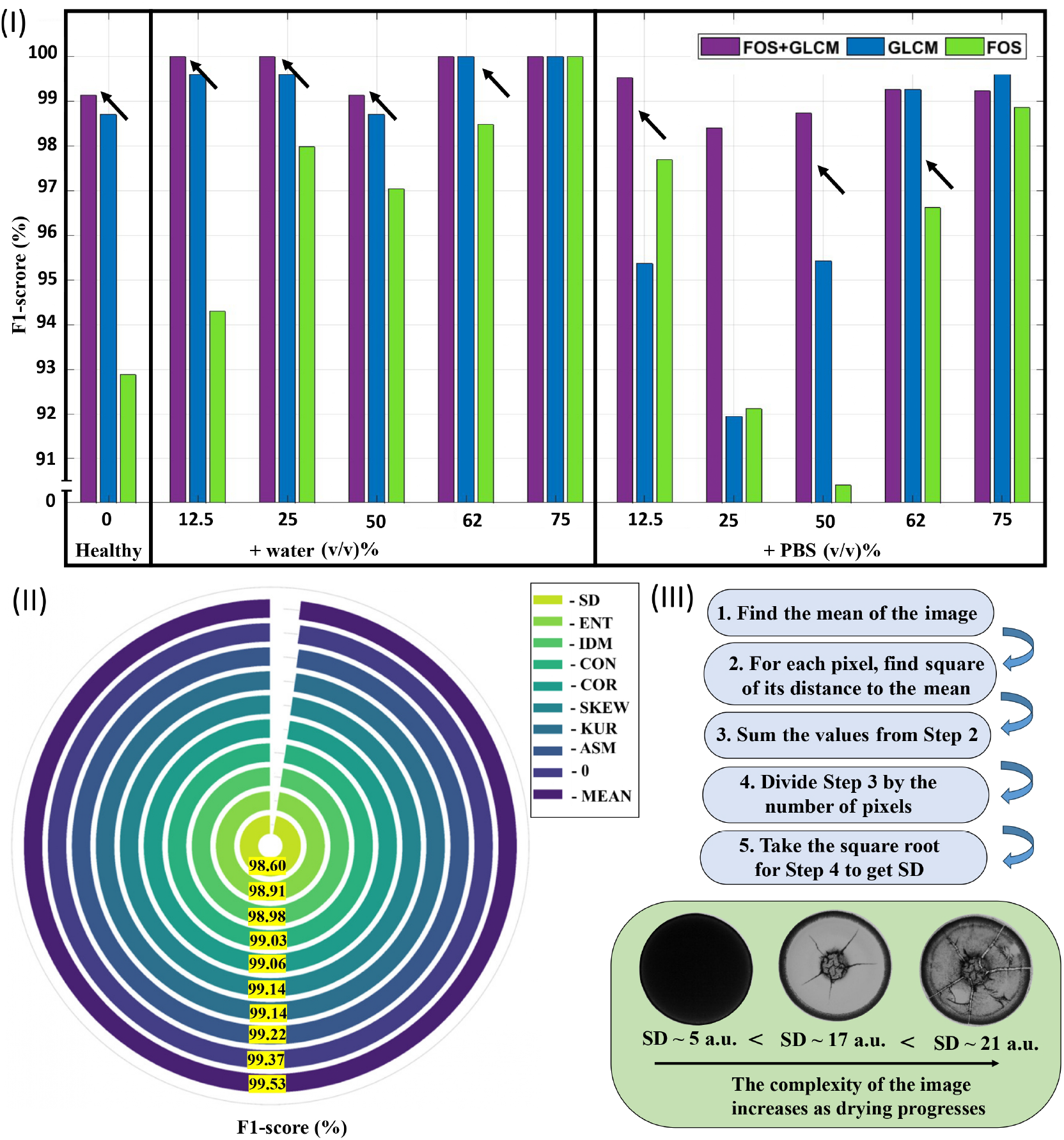
(I) Bar plots presenting F1-scores (in percentage) that offer a comparative analysis of random forest (RF) performance across different feature sets: (a) First-order statistics (FOS), (b) Gray-level co-occurrence matrix (GLCM), and (c) Combined FOS+GLCM. (II) Radial plots illustrate the average F1-score (in percentage) when individual features are omitted during RF implementation. Here, excluded features are denoted as ‘-feature’. The notation ‘-0’ signifies the inclusion of all features. The abbreviations SD, ENT, IDM, CON, COR, SKEW, KUR, and ASM correspond to standard deviation, entropy, inverse difference moment, contrast, correlation, skewness, kurtosis, and angular second moment, respectively. (III) Algorithmic depiction of SD computation of an image, highlighting the correlation between image SD and the system’s complexity as drying progresses. The SD of the droplet is directly linked to the escalating complexity, with values ascending from ∼5 to ∼21 arbitrary units (a.u.).

The initial image is captured approximately 50 − 90 seconds after droplet deposition, corresponding to around 0.06 − 0.08*t*_*f*_ [refer to the bottom panel of Fig. 2(I-II)]. Initially, these images display a uniform dark texture. During this stage, droplet height and contact angle decrease [33]. The dominance of outward capillary flow over surface tension-induced Marangoni flow leads to the movement of blood particles toward the periphery of the droplet. In the subsequent middle stage, there is a shift in the fluids front from the periphery toward the center of the droplet. This behavior is consistent with the drying process observed in other bio-colloids [16, 17]. As water evaporates and the droplet remains pinned to the substrate, mechanical stress accumulates. To relieve this accumulated stress, cracks start to initiate and propagate within the droplet. These cracks can appear in both radial and orthoradial directions. The middle panel of Fig. 2(I) visually represents these cracks forming within the droplet. The initiation and propagation of cracks can result from the complex interplay between drying-induced stresses, capillary forces, and the biophysical properties of the drying blood droplet. In the final stage, distinct patterns manifest as image textures change both spatially and temporally. For instance, at 12.5% (v/v), an inhomogeneous texture appears at the central region (visible at 0.71*t*_*f*_), eventually covering the entire droplet by the end of the drying process. When comparing textures of blood droplets at 12.5 and 75% (v/v), the level of inhomogeneity differs significantly. While 12.5% (v/v) exhibits dark and light gray patches, 75% (v/v) displays an overall lighter gray shade. Additionally, the central region expands as concentration increases from 12.5 to 75% (v/v). The type of cracks also varies: 12.5% (v/v) presents large radial and chaotic cracks in the central region, while 75% (v/v) features fewer radial cracks around the peripheral ring and none in the central region [refer to the top panel of 12.5 to 75% (v/v) in Fig. 2(I)].

Blood droplets containing PBS exhibit behaviors akin to blood+water in the initial and first middle stages [refer to bottom and middle panels of Fig.2(II)]. However, the most interesting observations occur in the final stage of drying. At a concentration of 12.5% (v/v) of PBS, dendrite-like structures form in the central region of the droplet. In the case of blood droplets with concentrations ranging from 12.5 to 50% (v/v) of PBS, radial and orthoradial cracks develop. Surprisingly, no radial cracks are observed at a concentration of 62% (v/v) of PBS. Instead, a distinctive ring-like pattern emerges in the central region. Finally, at 75% (v/v) of blood+PBS, the central region of the droplet exhibits a heterogeneous and grainy texture without substantial cracks [refer to the top panel of Fig. 2(II)]. For 25 and 50% (v/v), entire droplets are light gray, with some dark textures at peripheral rings and crack linings. Textures become homogeneous and grainy for 62 and 75% (v/v).

Figure 2(III-IV) provides insight into the textural variation in blood droplets with and without abnormalities. The first-order statistics (FOS) encompass parameters like mean, standard deviation, skewness, and kurtosis of the droplet. On the other hand, the gray-level co-occurrence matrix (GLCM) incorporates features like angular second moment, contrast, correlation, inverse difference moment, and entropy. These textural statistics are extracted from time-lapse images captured during drying for different blood abnormalities (+ water and + PBS) and healthy blood (without any abnormalities). A distinctive pattern is evident in all these textural statistics as they vary with normalized time 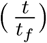. Notably, during the middle stage of the drying process, most of these statistical features exhibit dynamic behaviors characterized by peaks and dips, which are absent in the initial and final drying stages. The textural statistics presented in Figure 2(III-IV) vividly depict variations on both spatial and temporal scales. For instance, the temporal variation of the mean statistical feature of the droplet showcases a slow initial increase, followed by a rapid rise, a gradual decrease, and eventual saturation. On a spatial scale, during the initial drying stage, the mean values of blood droplets with 0 to 25%(v/v) concentration exhibit a range between 15 -25 a.u., whereas it’s 50-80 a.u. for droplets with 50 to 75%(v/v) concentration. In the final drying stage, the mean values range between 40 − 60 a.u. for 0 to 25%(v/v), 60 – 80 a.u. for 50 to 62%(v/v), and surpass 80 a.u. for droplets with 12.5% (v/v) concentration.

The FOS and GLCM statistics observed for blood+PBS [Figure 2(IV)] exhibit similar patterns capturing both temporal and spatial variations, comparable to those observed in blood+water droplets [Figure 2(III)]. However, the behavior of the droplets at both 0 and 75%(v/v) concentrations is distinct and noteworthy. These observations are consistent with previous findings in the realm of bio-colloids [45-46], indicating the broader applicability of the observed phenomena. Regardless of whether water or PBS is added, the drying process follows a consistent pattern in terms of its stages: an initial stage 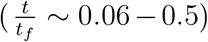, a middle stage (∼ 0.5 − 0.8), and a final stage (∼ 0.8 − 1.0). Interestingly, blood+PBS droplets maintain their dynamic behavior from the middle stage to the final stage 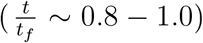, as depicted in Figure2(IV). In contrast, blood+water droplets exhibit their most dynamic behavior during the middle stage 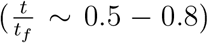 of the drying process, as illustrated in Figure 2(III). This suggests that the presence of salts in the PBS solution has a retarding effect on the process of pattern formation compared to the behavior observed when only water is present. Microstructural analysis through scanning electron microscopy provides additional insight into the distribution of blood components and PBS salts within the drying droplet setting. The morphology of the cellular components is also found to be changed due to different initial compositions. Further details regarding blood+PBS droplets with concentrations of 12.5 and 75% (v/v) are provided in Figures S2-S3 of the supplementary section.

The interesting point of this analysis is that the spatial and temporal dynamics of different image textures are able to capture the bio-physical changes within the blood droplets with respect to the different abnormalities. For instance, the initial stage is dominated by the flow and transport of the blood components, whereas the middle stage illustrates the mechanical stress induced by the drying process through the crack formation. Finally, the last stage depicts the aggregation of the blood components and provides us with clues about the dried patterns. It is also to note that the composition of the blood components with and without abnormalities are different and are well captured by the middle stage of the drying process. For instance, healthy blood contains cellular components with approximately 500 × 10^4^ of RBCs, 1 × 10^4^ of WBCs, and 40 × 10^4^ of platelets. A decrease of about a factor of 10 occurs when the abnormality of blood+water is at 75% (v.v). This suggests that these statistical features (both temporal and spatial) potentially carry information that allows for the classification of the blood samples.

### B. Machine learning (ML) for classifying different abnormalities in drying blood droplets

The steps to implement different MLs for classifying different abnormalities (types and concentrations) are detailed in the supplementary section.

#### 1. MLs for identifying different types of blood abnormalities

The normalized confusion matrix provides a comprehensive view of how well each ML performs in classifying the blood samples into healthy, blood+water, and blood+PBS categories. The performance evaluation includes (i) NB: The accuracy is around 58% for healthy blood, 39% for blood+water, and 87% for blood+PBS. The misclassifications are mainly between blood+PBS and healthy blood or blood+water. NB performs relatively poorly compared to other ML. (ii) SVM: After SVM replaces NB, the performance improves. The accuracy is around 74% for healthy blood, 84% for blood+water, and 100% for blood+PBS. SVM performs better than NB but still has room for improvement, especially in classifying blood+water. (iii) KNN: It performs better than both NB and SVM. It achieves accuracy ranging from 90% to 99% across all classes, demonstrating relatively minimal misclassification. (iv) DT: The accuracy is between 97% and 100% for all classes. DT performs well in accurately classifying the blood samples into their respective categories. (v) RF: It achieves similar accuracy as DT, ranging from 97% to 100%. RF shows the best performance, with minimal off-diagonal values in the normalized confusion matrix.

The average F1-score, a balanced metric considering precision and recall, is calculated for each ML. The hierarchy of performance based on F1-scores is: NB (63.7%) *<* SVM (90.8%) *<* KNN (97.8%) *<* DT (99.3%) *<* RF (99.7%). The F1-scores indicate that all ML can effectively differentiate between the blood samples with and without abnormalities using the extracted drying features. In fact, the different types of blood abnormalities, i.e., + water and + PBS, could also be identified. It’s important to note that these results are based on the testing data using a train-test split approach. To ensure the robustness of these MLs, 10-fold cross-validation is performed, which provides a more reliable assessment of their performance. The cross-validation results show that all ML algorithms maintain similar performance trends, with minor variations in accuracy scores. Both DT and RF demonstrate excellent performance, with accuracy above 99%, making them equally strong candidates for predicting the different blood sample categories. Therefore, while all the evaluated ML can differentiate between blood samples with and without abnormalities, DT and RF stand out as the best performers, offering accurate classification across the tested categories. Cross-validation reinforces their reliability and consistency in prediction [see Figure S4 in the supplementary section].

#### 2. Best feature selection and ML performance in classifying blood samples

Now that we have established the capability of ML to categorize these blood samples based on their FOS+GLCM statistical parameters, our next objective is to investigate whether all these features are necessary. This entails analyzing the effectiveness of utilizing FOS, GLCM, or their combination. To address this, we applied ML to diverse blood samples, which include healthy blood (0 (*v/v*)%), blood+water (12.5, 25, 50, 62, and 75 (*v/v*)%), and blood-PBS (12.5, 25, 50,62, and 75 (*v/v*)%). First-order statistics (FOS) comprise mean, standard deviation, kurtosis, and skewness. Additionally, we incorporated gray-level co-occurrence matrix (GLCM) parameters such as angular second moment, contrast, correlation, inverse difference moment, and entropy. We worked with eleven blood samples, considering four FOS features, five GLCM features, and nine features from the combined FOS+GLCM approach.

In Figure (I), the F1-scores are presented, comparing the performance of the RF when using FOS, GLCM, and FOS+GLCM features. Notably, employing GLCM features results in improved RF predictions for most blood samples, except for those at concentrations of 75 (*v/v*)% (+ water), 12.5, and 25 (*v/v*)% (+ PBS). Particularly, RF exhibits better predictive power when both FOS and GLCM features are combined instead of using them individually. Nonetheless, there are instances, such as at concentrations of 62 and 75 (*v/v*)% (regardless of + water and + PBS), where the distinction between GLCM and the combined FOS+GLCM features is less significant. On average, the F1-scores are ∼95% for FOS features, ∼97% for GLCM features, and ∼99% for the combined approach. This analysis suggests that either FOS or GLCM alone can effectively serve as feature vectors for RF implementation in this classification task. However, incorporating both FOS and GLCM as features enhances the F1-score. This improvement amounts to ∼2% compared to using GLCM alone and ∼4% compared to using FOS alone. Since the performance of any ML hinges on the quantity and nature of features, it’s crucial to ascertain whether RF s performance remains superior to other ML approaches, as seen in the previous case. In Figure S5 of the supplementary section, we showcase the average F1-scores for all traditional ML, including NB, SVM, KNN, DT, and RF, when applied to FOS, GLCM, and FOS+GLCM features. On average, all MLs exhibit enhanced classification performance when FOS+GLCM features are employed as the feature vector, whereas RF consistently outperforms the other ML.

To identify the key features crucial for this classification task, we executed an iterative procedure of excluding features and evaluating the RF classifier on the data collected during the drying process [see Figure (II)]. In the context of including all features without any exclusion, we denote this as - 0. Any omitted feature is represented as - feature. For instance, if the standard deviation (SD) is omitted, it’s indicated as - SD. When employing all features (- 0), the RF classifier achieves an F1-score of ∼99.3%. Upon removing the angular second moment (ASM) from the feature set, the RF’s performance remains largely unaffected, yielding an F1-score of ∼99.2%. Similar resilience in performance is observed when excluding Skewness (- Skew) or Kurtosis (- Kur) from the feature set, resulting in F1-scores of ∼99.1% for both cases. Similarly, this trend holds for - COR (removing correlation) and - CON (removing contrast), yielding an F1-score of ∼99.0%.

Intriguingly, when the Mean is removed from the feature set, the RF classifier exhibits improved performance compared to using all features together. This enhancement corresponds to a ∼0.16% increase in performance when comparing - 0 and - MEAN configurations. This counterintuitive observation suggests a negative contribution of the Mean feature to the classification task. This could be attributed to Mean values not effectively capturing finer nuances, displaying a consistent trend across nearly all blood samples [see Figure 2(III)-(IV)], unlike the other features. Upon excluding the inverse difference moment (IDM) from the feature set, the RF s F1-score drops to ∼98.98%, indicating a decrease of ∼0.40%. Eliminating the entropy feature (-ENT) results in an F1-score of ∼98.91%, indicating a ∼0.45% decline compared to - 0 configuration. However, when the SD is removed from the feature set, the RF s performance decreases significantly to ∼98.60%, resulting in a decline of ∼0.76% based on the F1-score. This suggests that SD contributes significantly to the overall feature set. In summary, the hierarchy for feature importance can be delineated as follows: SD *>* entropy *>* IDM *>* contrast *>* correlation *>* kurtosis = skewness *>* ASM *>* mean. Nonetheless, the analysis also implies that SD, entropy, and IDM share equal importance, as their absence reduces approximately ∼1% in RF s performance.

It’s important to recognize that GLCM captures spatial relationships among pixels using second-order statistics within a defined neighborhood, while FOS employs histograms to represent statistical feature distributions. The mathematical definitions of each feature are described in the supplementary section. Within FOS, mean and SD are straightforward to comprehend. Mean represents the average gray level intensity value within the Region of Interest (ROI), while SD quantifies the extent of variation around this mean. Skewness gauges the asymmetry of the pixel value distribution around the mean, and Kurtosis measures the distribution’s peakiness within the ROI. A higher kurtosis indicates a distribution with greater mass towards the histogram s tails [1,47].

For a better understanding, an algorithmic illustration is displayed of how SD is determined for a droplet (ROI), and how SD correlates directly with system complexity [see Figure (III)]. Low complexity or low heterogeneity is translated as low SD or entropy value (indicating high homogeneity). It is depicted in the initial stage of the drying process [see Figure 2(III-IV)], or the first image of Figure (III) indicates an ROI with a limited range of gray level values. Conversely, high complexity or high heterogeneity is reflected as a high SD or entropy value (indicating low homogeneity). It is shown in the final stage of the drying process [see Figure 2(III-IV)], implying significant variations and patterns within the ROI of the last image in Figure (III) (similar to what is observed in other systems, see [47]).

Additionally, IDM values are influenced by the image s dynamics-a low IDM value arises from static variations in the ROI, while a relatively higher value arises when substantial dynamics occur, potentially peaking in the middle stage of the drying process [see Figure 2(III-IV)]. Meanwhile, ASM gauges ROI homogeneity, yielding low values for a homogenous ROI with few gray levels. Contrast emphasizes local intensity variation, while correlation quantifies pixel correlation across the entire ROI[1]. Consequently, it becomes evident that the mean, kurtosis, skewness (from FOS statistics), ASM, contrast, and correlation (from GLCM statistics) alone lack the depth required for traditional ML to classify distinct blood samples effectively. Contrastingly, SD (from FOS statistics), IDM, and entropy (from GLCM statistics) prove pivotal for ML to distinguish between various blood samples. This underscores the importance of these features in capturing the necessary information for accurate classification.

#### 3. Performance of Neural Network in classifying blood samples

To assess whether comparable performance can be achieved using neural networks (NN) to classify these blood samples, we have adopted two different approaches: traditional ML and NN. The inputs consist of images, and a convolution neural network (CNN) is directly employed. Simultaneously, textural features (FOS and GLCM) are extracted and used in RF and artificial neural networks (ANN). The input, analysis, and goal to predict the eleven blood sample classes are shown in Figure 6(I).

**FIG. 6.**
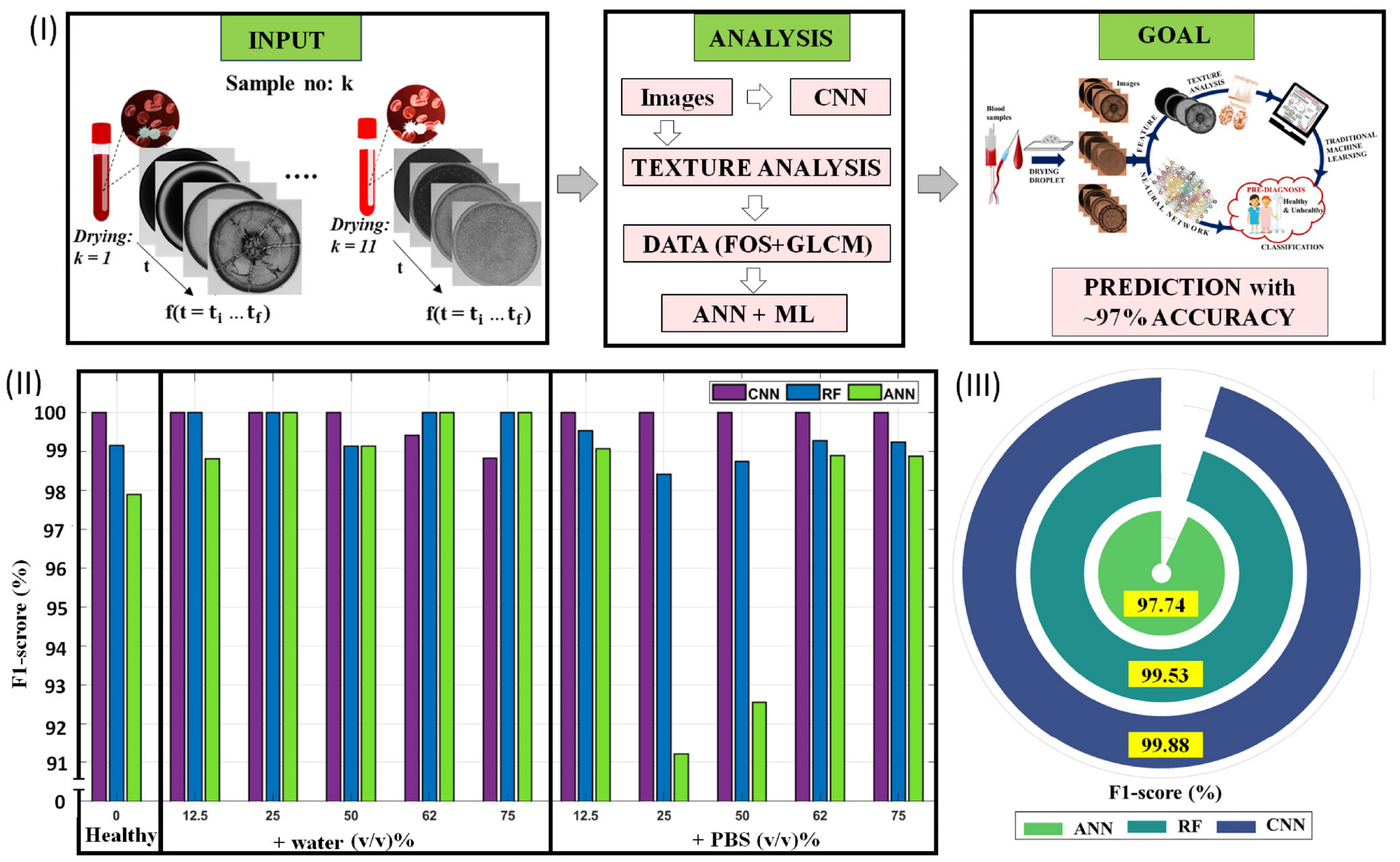
(I) Illustration of two distinct approaches: Approach 1: Direct utilization of images as inputs into a convolution neural network (CNN). Approach 2: Leveraging traditional ML, employing random forest (RF) and artificial neural networks (ANN). These models are based on the first-order statistics (FOS) and gray-level co-occurrence matrix (GLCM) features extracted from images using image processing techniques. The objective is to predict classes encompassing various blood compositions: healthy blood (0 (*v/v*)%), blood+water (12.5, 25, 50, 62, and 75 (*v/v*)%), and blood+PBS (12.5, 25, 50, 62, and 75 (*v/v*)%). (II) Bar plots showcasing the performance evaluation of CNN, RF, and ANN for each class. (III) Radial Plot illustrating the average F1-score (in percentage) across all classes.

The performance evaluation of CNN, RF, and ANN based on the F1-score is depicted in Figure 6(II-III). However, before delving into this, it s crucial to ascertain the presence of underfitting or overfitting issues in these NN implementations. Like other traditional ML, ANN and CNN are trained on a training dataset. Its suitability can be gauged by its performance on validation (testing) dataset-data that has not been encountered during training. Figures S6-S7 in the supplementary section illustrate the loss and accuracy curves as functions of epochs. The loss curve reflects the reduction of noise signals during training, and its progression is smooth without noticeable spikes or fluctuations. Accuracy also tends to saturate as epochs increase. The convergence of training and validation curves without divergence indicates the absence of underfitting or overfitting issues. This suggests that these neural networks can predict the eleven blood sample classes.

The normalized confusion matrix for CNN, RF, and ANN is presented in Figure S8 of the supplementary section. These results indicate that CNN misclassifies only one class with ∼1% error. Notably, ∼99% of the 75 (*v/v*)% blood+water samples are accurately predicted, with 1% being misclassified with the 12.5 (*v/v*)% blood+PBS. RF exhibits misclassifications in predicting healthy blood [0 (*v/v*)%], 25, and 62 (*v/v*)% blood+PBS. Interestingly, ANN performs worse than CNN and RF, with misclassifications in healthy blood, 62 (*v/v*)% blood+water, and 25, 50, and 75 (*v/v*)% blood+PBS. This pattern is consistent when the F1-scores for each class are visualized in a bar plot in Figure 6(II). The hierarchy CNN *>* RF *>* ANN in predicting blood samples is upheld in most scenarios. However, in the case of blood-water, both RF and CNN or RF and ANN perform similarly. Consequently, it s challenging to definitively state which approach is superior for this classification task [illustrated by the average F1-score in the radial plot in Figure 6(III)]. We can infer that both traditional and neural network ML approaches are equally effective for predicting blood samples, yielding F1-scores of over ∼97%.

### C. Competitiveness of the proposed method compared to the state of the art

The current landscape of healthcare systems calls for two critical aspects: firstly, gaining insights into the biophysical and biochemical properties of fluids to foster a fundamental and quantitative understanding related to patient health, and secondly, leveraging these insights in conjunction with image recognition techniques to develop simplified diagnostic or screening methods capable of detecting diseases at an early stage. This foresees a potentially life-saving innovation that enables swift and efficient diagnoses in critical scenarios. However, disease diagnosis in the context of drying droplet patterns poses a significant challenge due to the diversity of diseases and human samples involved.

Table I offers a comprehensive assessment of various MLs applied to diverse colloidal systems in drying droplet configurations. This table underscores the competitiveness of the proposed method when compared to state-of-the-art approaches. The investigated systems encompass bio-fluids, tap water, milk, and more. Notably, blood samples are correlated with different physiological states of cyclists [39], and cerebrospinal fluids samples are linked to healthy individuals and Alzheimer s Disease (AD) patients [50]. Their dried patterns are classified using Principal Component Analysis (PCA), an unsupervised ML. Additionally, peptides (A*β*) associated with AD are classified based on their primary and secondary structures using transfer learning (NasNet-Large) from dried droplets, achieving 99% accuracy [51]. Similarly, transfer learning, ResNet is used on the images acquired from the dried droplets of blood and urine, differentiating healthy and patients suffering from bladder cancer [49].

**TABLE I.**
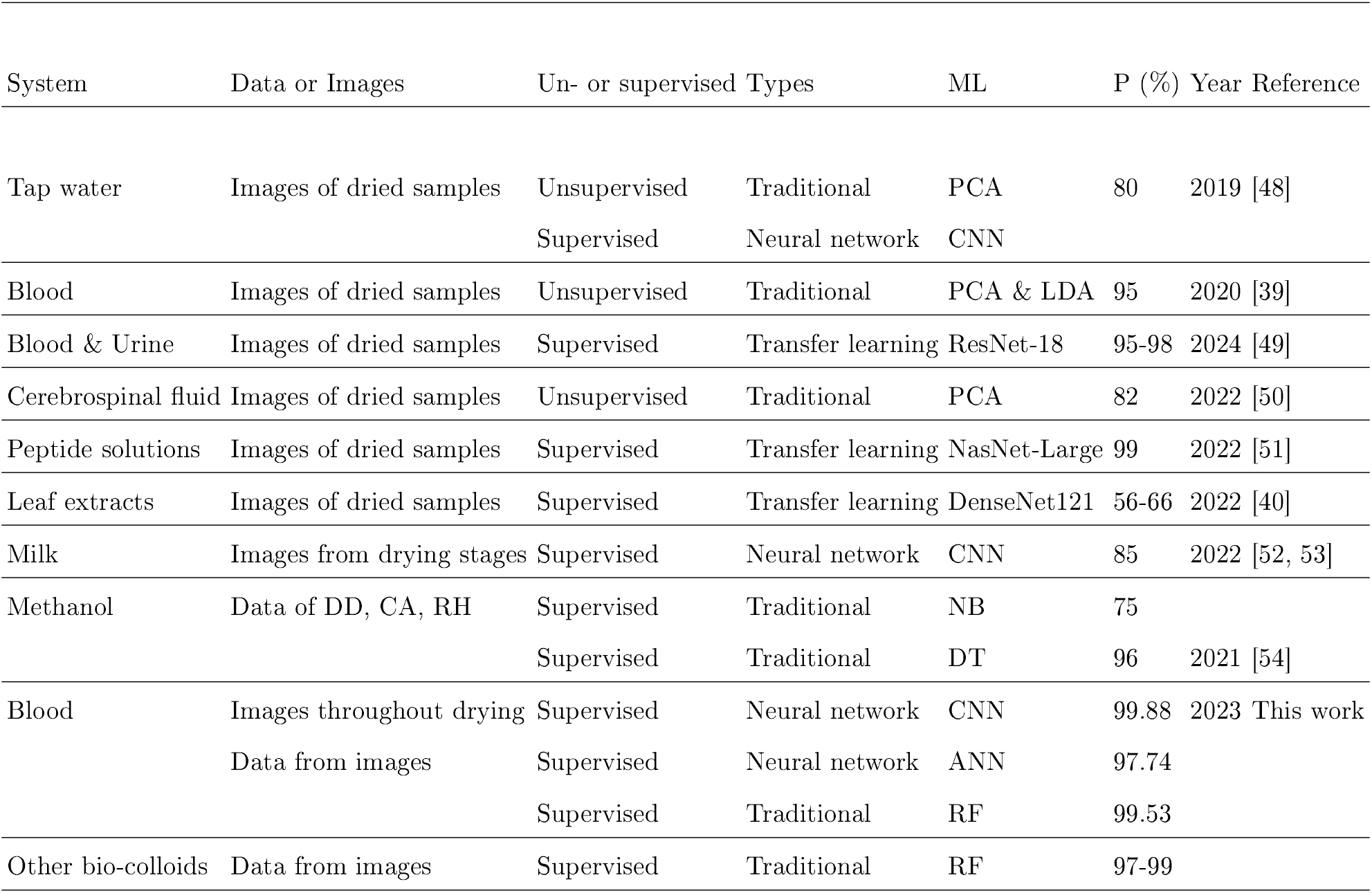
Summary of recent studies employing diverse machine learning (IL) techniques to classify patterns observed in drying droplet configurations. Abbreviations used include P: Performance, PCA: Principal Component Analysis, CNN: Convolution Neural Network, LDA: Linear Discriminant Analysis, NB: Naive Bayes, DT: Decision Trees, RF: Random Forest, ANN: Artificial Neural Network, DD: Droplet Diameter, CA: Contact Angle, RH: Relative Humidity. It provides insights into the efficacy and applicability of these IL approaches in analyzing complex patterns.

Other examples include milk, where contamination, coagulation, and decay are identified [52,53] and tap water. The diverse fingerprints of these dried droplet patterns are identified using CNN [48]. The plant extracts in which different textures are classified using transfer learning (DenseNet-a densely connected pre-trained CNN) [40]. Traditional ML approaches (DT and NB) are used in methanol drying droplets to estimate drying stages using data related to droplet base diameter, contact angle, and relative humidity [54]. Importantly, all these studies (except[54]) involve (i) the acquisition of images via optical microscopy and (ii) the implementation of neural networks and PCA on these acquired images.

The illustration of the drying droplets, pattern recognition, and the application of ML with respect to the existing literature is exhibited in Figure 7(I-II). We also explore varied avenues for incorporating ML techniques [indicated by the gray arrows], wherein images are transformed into analyzable data through image processing techniques. The textural attributes, specifically first-order statistics (FOS) and gray-level co-occurrence matrix (GLCM), serve as fundamental features. These features are then subjected to different traditional ML, [such as Decision Tree (DT), Random Forest (RF), Support Vector Machine (SVM), K-Nearest Neighbors (KNN), and Naïve Bayes (NB)] using the data derived from textural analysis. This study presents a proof-of-concept, demonstrating an impressive accuracy range of 97 − 99%, by synergizing simple drying experiments with ML methodologies to investigate evolving patterns during the drying process of human blood containing different types of abnormalities. Consequently, this study answers that drying pattern dynamics and ML effectively differentiate between blood samples with and without abnormalities.

**FIG. 7.**
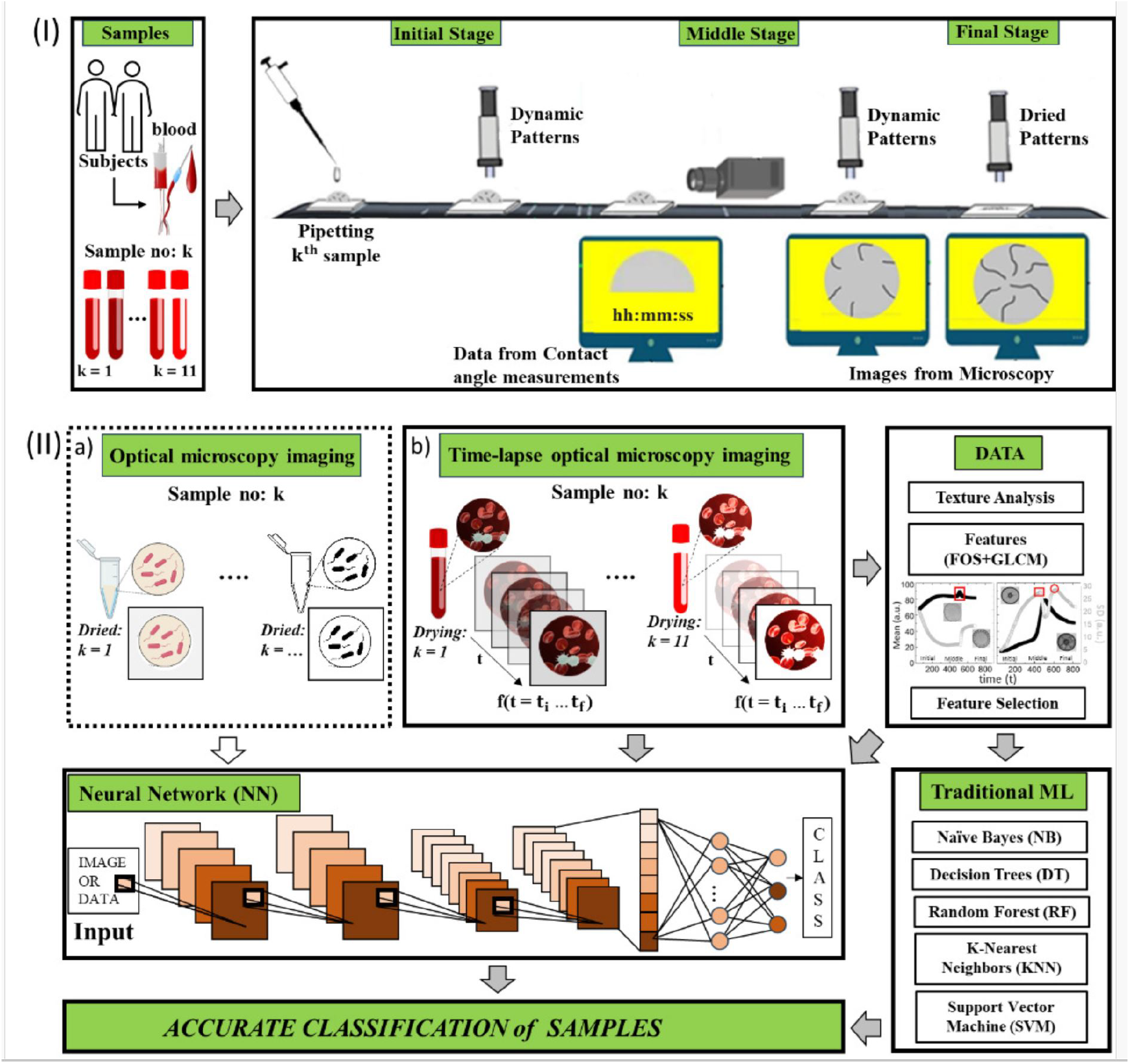
(I) Comparative overview of drying experiments between our proposed method and previous literature. Our approach captures the entire drying evolution, spanning from droplet deposition to the fully dried states. In contrast, existing literature either utilizes data from the contact angle setup or images from optical microscopy at specific stages. (II)(a)-(b) Our technique employs time-lapse images, capturing the dynamic evolution, while prior studies depict images at isolated stages (middle or final) of the drying process. The depiction of the ML implementation is denoted by white (representing prior literature) and gray (indicating our approach) arrows. Our proposed methodology converts images into data, employs image processing techniques for quantifying temporal textural parameters [first-order statistics (FOS) and gray-level co-occurrence matrix (GLCM)], and subsequently selects pertinent features. Various traditional ML are utilized [Decision Tree (DT), Random Forest (RF), Support Vector Machine (SVM), K-Nearest Neighbors (KNN), and Na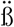ve Bayes (NB)). In contrast to previous approaches, our method competes by implementing traditional ML on data and employing Neural Networks (NN) separately for images and data.

Furthermore, in this current study, the fusion of spatial and temporal features during droplet drying is shown. It offers several advantages: (i) Enhanced precision in classifying these patterns; (ii) Eliminating the time-consuming and labor-intensive task of depositing 20-50 droplets to build image datasets for different diseases; and (iii) Overcoming the challenge of obtaining a sufficient volume of diseased blood for making 20-50 droplets, which is significantly more than the 3-5 droplets required. Notably, the time-lapse images captured at two-second intervals can swiftly generate 400-600 images for the entire drying process of a droplet with a radius ∼1 mm, volume of ∼1 *μ*L, dried at ambient temperature of about 25°C and RH ∼ 50%. Evaluating the performance of each ML, this study stands as the first to employ (i) images acquired during the drying process, (ii) data extraction using textural statistics (FOS and GLCM) closely related to different drying process events, (iii) use of both images and data in traditional and NN-based ML, and (iv) achieve a high performance exceeding 97%. However, neural network-based ML typically demands substantial data, high GPU capabilities, and advanced hardware, whereas, for low-resource scenarios, RF can be employed as a traditional ML using textural features.

The approach of utilizing these textural features within a drying droplet context is advantageous, as these features are unique and are directly related to the variations in blood compositions, offering biophysical insights to be used as a futuristic quantitative tool in implementing any data-driven approaches.

To ascertain the versatility of this approach, RF is implemented on other bio-colloidal systems using FOS+GLCM textural statistics, yielding F1-scores of 97 − 99% (see Figure S9 and Table S1 in the supplementary section). It’s imperative to compare the performance of each ML method while applied to other bio-colloidal systems, as the efficacy of traditional or NN-based ML hinges on data quality and quantity, as well as feature vector composition. Prior approaches may have struggled to translate effectively due to the necessity of involving multi-scale experimental tools, image processing techniques, machine learning, and connecting with different bio-fluids abnormalities or diseases in patients. Thus, this study aims to establish an interdisciplinary approach to bio-mimetic drying droplets, where abnormalities are simulated by adding water and phosphate buffer saline and progress toward realizing a diagnostic marker.

## IV. CONCLUSIONS

In this study, we show how the initial composition of bio-fluids influences the dynamics of pattern formation during the drying process. The composition of the blood samples is varied by adding water and buffer, and the time-lapse images are captured using optical microscopy. An efficient classification method is proposed by analyzing these evolving drying patterns rather than the dried patterns that have mostly been done so far. In addition, the highlight of the paper is the quantitative image analysis, where the textural statistics are derived from the time-lapse images, capturing the morphology differences among different blood abnormalities. Five different traditional machine learning and artificial neural networks were implemented on the textural data, whereas convolution neural networks were directly applied to the images. An accuracy of more than 97 % is achieved in classifying eleven different blood samples. This means that the time series of the drying process is essential, not just the traditional final dried state. An additional benefit of using this time series is that the amount of solution used in the experiment can be minimized. Bio-fluids such as blood, plasma, urine, and saliva have significant individual differences and are not suitable for mixing samples from multiple patients. This method, i.e., drying pattern dynamics and machine learning, therefore, represents a revolutionary advance in the potential for disease diagnosis to identify and classify diseases from small amounts of raw bio-fluids obtained from a single patient, advancing the fields of data science, biotechnology, and colloidal physics improving public health.

## Supporting information

This is the supplementary file.

## Data Availability

All data produced in the present study are available upon reasonable request to the authors.

## ACKNOWLEDGMENTS

This work is supported by the Graduate School of Arts and Sciences at The University of Tokyo, Japan. A.Pal would like to express her gratitude to the Tinkerbox community at Worcester Polytechnic Institute (WPI), USA, for providing an opportunity to initiate the research of bio-fluids. This research receives funding from UTOPIA (supported by AMED under Grant Number JP223fa627001), and the Japan Society for Promotion of Science (JSPS), KAKENHI Grant No. 23KF0104. A. Pal expresses appreciation for the JSPS International Postdoctoral Fellowship for Research in Japan (Standard) for the period 2023-25. This work is going through a patent application at the US Patent Office. The inventors of this patent are A. Pal and M. Yanagisawa. The application number is 63/ 566578. The patent was filed on the 22nd of February 2024.

## AUTHOR CONTRIBUTIONS STATEMENT

A.P. and A.G. designed and conceptualized the research. A.P. performed the experiments and analyzed the data. M.Y. helped A.P. in organizing and validating the results. A.G. validates the machine learning results. A.P. wrote the initial draft of the paper. A.P., M.Y., and A.G. revised the final draft. All authors have given approval to the final version of the manuscript.

## ETHICAL STATEMENTS

Approval for this study was not required because the blood samples were purchased from a reputable company that adheres to all applicable rules and regulations regarding the collection and handling of biological specimens. All procedures followed ethical guidelines to ensure the integrity and ethical conduct of the research.

## DATA AVAILABILITY STATEMENT

The data that support the findings of this study are available from the corresponding author upon reasonable request.

