## Supplementary material for "Time-Lapse Quantitative Analysis of Drying Patterns and Machine Learning for Classifying Abnormalities in Sessile Blood Droplets": This is the supplementary file.

#### I. MATERIALS AND EXPERIMENTAL METHODS

##### A. Image processing

The image's texture is quantified by extracting the different parameters. The image is a grid of pixels. The first-order statistics (FOS) is based on the gray level distribution of the pixel values without intervening in the interpixel relationships. The Gray Level Co-occurrence Matrix (GLCM) are the second-order statistical parameters that are calculated from the spatial relationship between two neighboring pixels [1]. The mean ($I$) and the standard deviation ($SD$), skewness ($SKEW$), and kurtosis ($KUR$) are calculated for the droplet region from the time-lapse images. Mathematically, these parameters can be defined as follows; where, $I(i,j)$ is the gray scale value of the pixel at location $(i,j)$ in the image, and $N$ is the number of pixels in the image [2].

$$I = \frac{1}{N}\sum_{i,j=0}^{N} I(i,j) \tag{1}$$

$$SD = \sqrt{\frac{\sum_{i,j=0}^{N}(I(i,j) - Mean)^2}{N-1}} \tag{2}$$

$$SKEW = \frac{\sum_{i,j=0}^{N}(I(i,j) - Mean)^3}{(SD)^3} \tag{3}$$

$$KUR = \frac{\sum_{i,j=0}^{N}(I(i,j) - Mean)^4}{(SD)^4} \tag{4}$$

For the GLCM parameters, we have considered only the horizontal (0°) orientations and pixel displacement as 1. The five parameters are extracted that include the angular second moment ($ASM$), correlation ($COR$), contrast ($CON$) inverse difference moment ($IDM$), and entropy ($ENT$). We assume the reference pixel as $i_c, j_c$, and the displacement $d$ as the distance from that reference. The direction is determined by the $\theta$ as $D_x = Dcos\theta$ and $D_y = Dsin\theta$. If the number of occurrences at location $(i,j)$ at particular $D$ and $\theta$ is $C_D(i,j)$, and $N$ is the number of pixels in the image, then the probability for the changes occurring between $i$ and $j$ at a particular $D$ and $\theta$ is $p_D(i,j)$. Then, the $p_D(i,j)$ is defined as $\frac{C_D(i,j)}{\sum_{i,j=0}^{N} C_D(i,j)}$.

*

$$ASM = \sum_{i,j=0}^{N} p_D(i,j)^2 \tag{5}$$

$$CON = \sum_{i,j=0}^{N} (i-j)^2 p_D(i,j) \tag{6}$$

$$COR = \sum_{i,j=0}^{N} \frac{(i-\mu_x)(j-\mu_y)p_D(i,j)}{\sigma_x \sigma_y} \tag{7}$$

where, $(\mu_x, \mu_y)$ and $(\sigma_x, \sigma_y)$ are the means and the standard deviations of $(p_D(x), p_D(y))$, respectively.

$$IDM = \sum_{i,j=0}^{N} \frac{p_D(i,j)}{1+|i-j|} \tag{8}$$

$$ENT = -\sum_{i,j=0}^{N} p_D(i,j)\ log\ p_D(i,j) \tag{9}$$

#### B. Machine learning (ML)

##### 1. *Traditional ML*

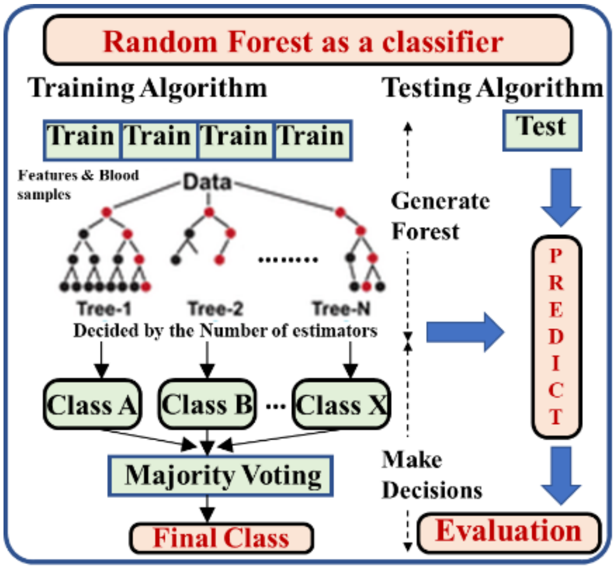


FIG. S1. The architecture of the Random Forest (RF) is shown for classifying different blood samples (classes) using the extracted parameters (FOS+GLCM) as features.

##### 2. *Neural Networks (NN)*

Convolution neural networks (CNN): the images are resized as $150 \times 150$. The three channels of the images are considered in this study. The layer added includes the *Conv2D()* with two different filters, 64 and 128, and the *kernel_size = (3, 3)*. The *activation = 'relu'*, and the *L2 regularization* of *kernel_regularizer=regularizers.l2* with the learning rate of *l=0.01* is also used. The *MaxPooling2D()* with the *pool_size = (2,2)* is also added. Prior to the another convolution and activation layers of *Conv2D(64, (3, 3), activation = 'relu')*, we used the Dropout layer of 40%. The *MaxPooling2D()* with the *pool_size = (2,2)* is again added, with the Dropout layer of 40%. Finally, we change the dimensions by *Flatten()*. We added a dense layer of filter size 64 and activated it with the RELU function. The 20% of the connected neurons are again dropped out. The output layer has a dense layer of size 11 corresponding to the number of classes and is activated using the *Softmax* function. This neural network is performed for the number of *epochs = 10* and *batch sizes = 16*. Note that the "relu" is the Rectified Linear Unit (ReLU), a non-linear function used to excite the layers. It provides an output only when the input value is non-zero numeric.

Artificial neural networks (ANN): the three Dense layers were added of three different filters, 512, 256, and 64. For each layer, the *activation = 'relu'*, and the *L2 regularization* of *kernel_regularizer=regularizers.l2* with the learning rate of *l=0.001* are used. The Dropout layer of 30% is added to each layer. Finally, the output layer has a dense layer of size 11 corresponding to the number of classes and is activated using the *Softmax* function. This neural network is performed for the number of *epochs = 100* and *batch sizes = 32*.

### II. RESULTS AND DISCUSSIONS

#### A. Visual inspection and textural analysis of the blood drying droplets

##### 1. *Drying evolution of healthy blood*

Fig. S2 shows the time-lapse images of the drying evolution of the human whole blood droplet. The details can be found in [3]. However, to compare the droplets with and without added phosphate buffer saline (PBS), we describe each event in the healthy blood droplet during the drying process. The original images of $3664 \times 2748$ pixels were cropped to

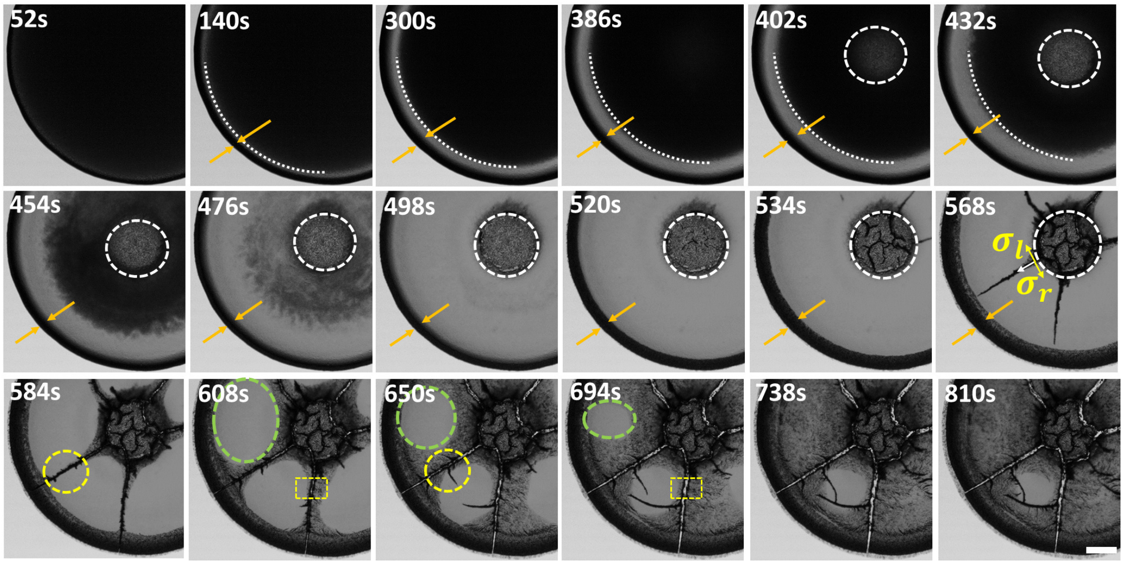


FIG. S2. Time-lapse images of the (healthy) blood droplet during drying. These displayed images are cropped from the original ones for better visualization. The top panel illustrates the formation of the peripheral band (depicted with orange arrows) and the movement of the fluid front. The white dashed arc line shows that the front moves uniformly from the periphery of the droplet up to 432s. The texture in the central region becomes lighter (outlined with a white dashed circle) and separates it from the central region. The middle panel captures images from 454s to 568s. It describes the non-uniform movement of the front, the appearance of the random cracks in the central region, and the initiation of the radial cracks from the transition of the central and the peripheral regions. The white arrows sketch the propagation of the radial cracks. The $\sigma_l$ and $\sigma_r$ demonstrate the symmetric stress fields of these cracks (depicted with yellow arrows). The bottom panel records the crack propagation and the texture change from light to dark gray from 584s to 810s. The yellow dashed circles show the origin of the micro flaws around the radial crack, whereas the yellow dashed squares portray the widening of these radial cracks. The green dotted oval line shows the texture change in the domains created by these radial cracks. The time ($t$ in seconds) during the drying process is exhibited in the top-left of each image. The scale bar in the bottom-right at 810s corresponds to 0.2 $mm$.

$1908 \times 1902$ pixels and presented for clear visualization. The first image is captured at $\sim$50s after the droplet deposition on the coverslip (substrate). The droplet exhibits a uniform dark texture; no visible change is observed initially. As time progresses, a fluid front moves uniformly from the periphery towards the center from ∼140 to ∼430s. The white dashed arc line shows this uniform movement in the top panel of Fig. S2. As the front proceeds, a smooth gray texture behind it starts appearing. This results in a dark peripheral band (depicted with orange arrows) from ∼140s. While the gray texture appears with this front movement, the dark texture around the center of the droplet (marked with a white dashed circular line) turns lighter from ∼400s.

The next stage begins with the non-uniform movement of the fluid front. It completely disappears within ∼500s, shown in the middle panel of Fig. S2. Once the front vanishes, it becomes clear that there exist two regions. The white dashed circular line helps us to identify these two regions. While the texture is light gray, the central region is slightly darker up to ∼570s. In the meantime, there is an increase in the width of the peripheral band. This can be prominently noticed when the snapshots at 300s and 534s are compared. As the drying process evolves, some cracks appear in the central region (image at 520s). These cracks are random and are observed to grow till ∼570s. Furthermore, some cracks initiate from the transition region (between the peripheral and the central regions) at ∼530s. Unlike the cracks in the central region, these cracks propagate toward the periphery in a particular direction. The white arrows indicate that the crack propagation is radial. The symmetric stress fields ($\sigma_l$ and $\sigma_r$) of these cracks are described with the yellow arrows. Due to these radial cracks, the peripheral region gets divided into large-sized domains, as seen in the snapshot at 568s. The keen observation of the bottom panel (the time snapshots from t $= 584 - 810$s, makes it clear that the dried droplet is no longer is firmly attached to the substrate and slightly displaced from the original position compared to that of the initial stage of the drying process.

##### *2. Drying evolution of blood with PBS at 12.5(v/v)%*

Fig. S3 shows the time-lapse images of the drying evolution of the blood droplet with added PBS at 12.5% (v/v). These images were cropped from the original images to $1812 \times 1656$ pixels. At the initial stage of the drying process, the blood + PBS droplet behaves similarly to that of the whole blood (see Fig. S2). The formation of the peripheral band, the uniform movement of the fluid front, and the appearance of the central light gray texture occur for the time duration of 55s and 467s, shown in the top panel of Fig. S3. All these

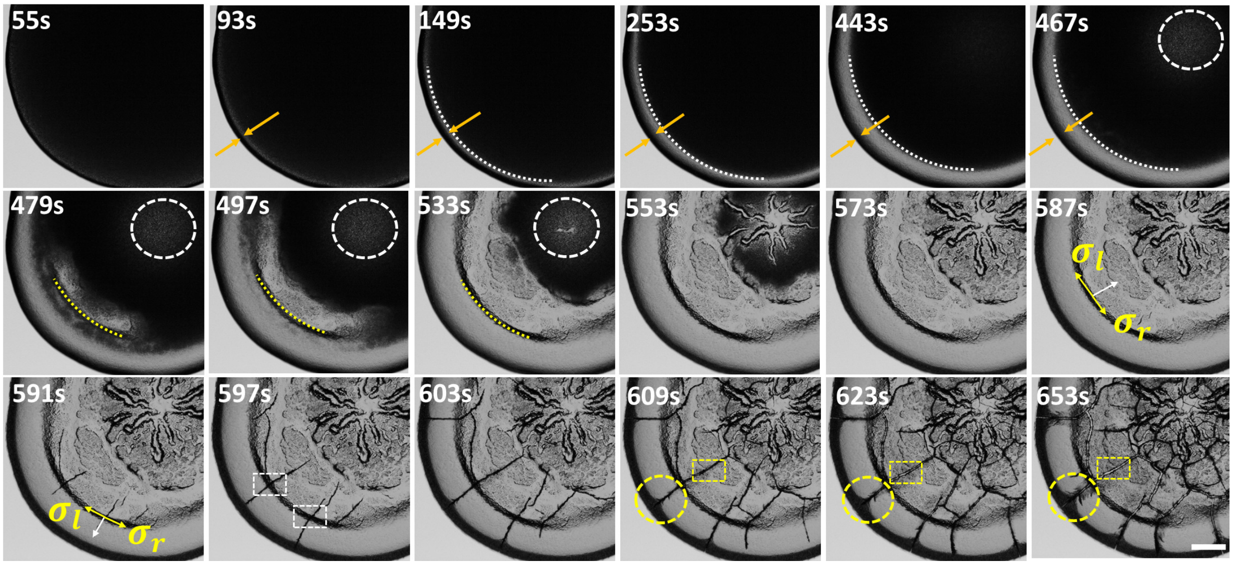


FIG. S3. Time-lapse images of the diluted blood droplets during the drying process at 12.5% (v/v). These displayed images are cropped from the original ones for better visualization. The top panel illustrates the formation of the peripheral band (depicted with orange arrows) and the movement of the fluid front. The white dashed arc line shows that the front moves uniformly from the periphery of the droplet up to 467s. The texture in the central region becomes lighter, outlined with a white dashed circle. The middle panel captures images from 479s to 587s. This panel emphasizes the non-uniform movement of the front and the appearance of the dendrite structure in the central region. The yellow dashed arc line separates the periphery from the central region. The white arrows sketch the propagation of the radial cracks. The $\sigma_l$ and $\sigma_r$ demonstrate the symmetric stress fields of these cracks (depicted with yellow arrows). The bottom panel records the crack propagation from 591s to 653s. The cracks grow in both directions at this transition, and a kink is spotted (white dashed rectangles). The yellow dashed circles display the origin of the micro flaws around the radial crack, whereas the yellow dashed rectangles portray the widening of these radial cracks. The time ($t$ in seconds) during the drying process is exhibited in the top-left of each image. The scale bar in the bottom-right at 653s corresponds to 0.2 $mm$.

events are found common in these droplets.

The next stage of the drying evolution begins with the non-uniform movement of the fluid front; however, the later events become different in both droplets. The peripheral and central regions can be identified. In contrast, in blood + PBS at 12.5%, a transition line appears (marked with a yellow dashed arc line in the snapshot captured at 479s) while the front still moves non-uniformly. This line continues developing until it fully grows into a ring. Finally, it separates the peripheral and the central regions similar to the healthy blood droplet (middle panel of Figs. S2 and S3). Interestingly a dendrite structure starts appearing in the center of the droplet at 12.5%. The growth of such a structure is evident for the time intervals between 533s and 573s. Its presence makes the drying evolution very different from that of healthy blood. The crack propagation initiates from the transition region at 587s. The white and the yellow arrows show the direction of the cracks and the symmetric stress fields, respectively (snapshots at 587s and 591s). Unlike the healthy blood-drying droplet, some cracks grow towards the central region, whereas some propagate towards the periphery. It is to be noted that the crack lines in the transition region are not straight. A kink around the transition line is spotted (white dashed rectangles at the snapshot of 597s). As time advances, the cracks propagating towards the central region join the dendrite structure.

Akin to the healthy blood droplet, the branching and the widening of the radial cracks are found in blood + PBS at 12.5% from 609s to 653s. The yellow dashed circles display the origin of the micro flaws around the radial crack. In contrast, the yellow dashed rectangles portray the widening of these radial cracks in the snapshots captured from 609s. However, the texture of the crack domains does not become dark gray fully in the peripheral region. Furthermore, the droplet is observed to be in the same position throughout the drying process, unlike the healthy blood droplet (bottom panel of Figs. S2 and S3). Therefore, these visual inspection helps us to understand the minute details of the similarities and the dissimilarities of the events occurring during the drying process in the blood droplets with and without PBS.

Fig. S4(I-II) shows the different micro-structures in the blood droplet at $\phi$ of 75% (v/v). The smooth fragmented sheet in the corona region, the raptured structures in the peripheral cracks, the presence of the troughs and ridges in the transition region, and the presence of the hair-like structures in the crack edges are found at $\phi$ = 75% (v/v) [Fig. S4(II)]. Interestingly, the central region at $\phi$ of 75% (v/v) displays the crystal-like structures in addition to the ridges and the toughs. The cracks in the central regions are comparatively sharper. The regions within these cracks are more homogeneous. The energy-dispersive X-ray (EDX) analysis is exhibited in Fig. S4(III). The different symbols are presented to identify the specific area from where the counts are collected. A graph of the number of counts and

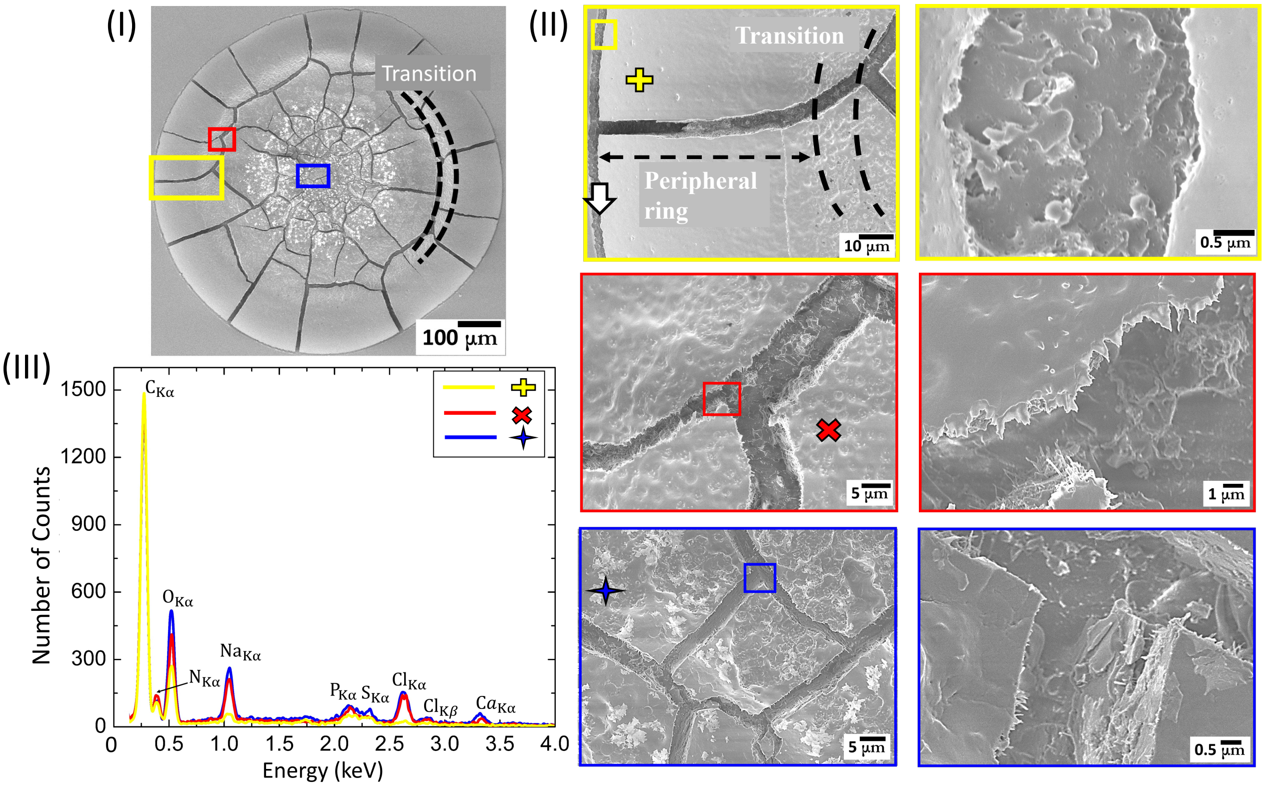


FIG. S4. The SEM images illustrating the microscopic structures at different length scales are shown in (I) and (II) at 12.5% (v/v). The top, middle, and bottom panels of (II), depicted with yellow, red, and blue-colored rectangles, detail the structures at the periphery and the corona, the transition of the corona and central regions, and the central region, respectively. The transition region is also specified with black dotted lines. An energy-dispersive X-ray (EDX) analysis consisting of the number of counts and the energy (keV) of different regions of the blood-dried droplet is shown in (III). The signals are symbolized and colored to match the area from where these were collected.

the energy (in keV) is plotted. The significant peaks are observed at 0.28, 0.53, 1.06, and 2.64 keV, characterizing the presence of the elements Carbon (C), Oxygen (O), Sodium (Na), and Chlorine (Cl), respectively.

The micro-structures at $\phi$ of 12.5% (v/v) are shown in Fig. S5(I-III). These structures are unique and very different from 75% (v/v). A transition region between the corona and the central regions is not observed here. Fig. S5(I-II) marks different regions of interest. The yellow-colored rectangle in the top panel of Fig. S5(II) details the structures at the periphery and the corona. The light and dark blue-colored rectangles in the middle and the bottom panels of Fig. S5(II) show different structures in the central regions. The corona region is smooth like what we observed at other $\phi$; however, it contains discoid-shaped aggregated structures. Almost no raptured structures are found within the cracks of the peripheral region. A closer view of the central region indicates the presence of different crystal-like structures [the middle and the bottom panels of Fig. S5(II)]. The dried droplet has no troughs and ridges at $\phi = 12.5\%$ (v/v). Similar to $\phi = 75\%$ (v/v), Fig. S5(III) also shows the significant presence of C, O, Na, and Cl elements. However, it is to be noted that the number of counts for the elements changes as we dilute the initial concentration of blood ($\phi$) from 75 to 12.5% (v/v). For example, the counts of C decreases from ∼1500 to ∼450 in the corona region for $\phi$ of 75 and 12.5% (v/v), respectively. It is found to be ∼300 for Na and Cl at $\phi$ of 75% (v/v) in the central region of the dried droplet. These counts of Na and Cl turn out to be more than ∼900 at $\phi = 12.5\%$ (v/v).

This section describes how the adulterant (1x PBS) affects the microscopic structures of the blood components (mainly RBCs, WBCs, and platelets) and the macroscopic crack patterns in the drying droplets. The whole blood [$\phi = 100\%$ (v/v)] is diluted by decreasing its volume and increasing the volume of PBS at a fixed concentration of 1x. This indicates that the $\phi$ of 75% (v/v) contains more number of blood components and less number of salts (ions) as compared to 12.5% (v/v). As the $\phi$ changes from 75 to 12.5% (v/v), the amount of the salts increases with the decreasing number of the blood components. The underlying physics of the drying process involving the blood components (RBCs, WBCs, and platelets) as we add the adulterants (DI and PBS) is detailed in [3]. In this article, we are mostly interested in the comparison of the blood+PBS dried deposits at $\phi = 75\%$ and 12.5% (v/v).

The dried droplet at 12.5% (v/v) looks very different from the whole blood [$\phi = 100\%$ (v/v)] or the diluted blood at 75% (v/v) [Figs. S1 in the supplementary section, **??**(I-II), S4(I-II), and S5(I-II)]. In terms of image texture, and microscopic structures, a distinction can be made at $\phi = 75\%$ and 12.5% (v/v). Some discoid shaped structures are found in the corona region at $\phi = 12.5\%$ $(v/v)$ [see Fig. S5(II)]. The EDX analysis show that the number of counts for Na or CL is quite low, suggesting that these are not salt residues, but the deformed cellular structures of the blood. This also indicates that the deformation of the blood components might have occurred due to two reasons (i) first one is the developing mechanical stress during the drying process and (ii) secondly, the change in the microenvironments due to the presence of excessive salts. No trough and ridges are found in the central region as we see at $\phi = 75\%$ $(v/v)$ [see Fig. S4(II)], which supports our previous observation in blood+DI [3]. The hairy structures that are present at the edge of the crack domains in the central region at $\phi = 75\%$ $(v/v)$ [see Fig. S4(II)], are also not observed, rather sharp crack lines are found at $\phi = 12.5\%$ $(v/v)$ [see Fig. S5(II)]. This indicates the reactivity or the activation of the platelets might be affected due to the presence of excessive salts. The reduction of the blood components at 12.5% (v/v) makes the dried droplet as a sea of salts [Fig. S5(I-III]. The SEM image exhibits a well-defined cubic structure in the central region, which is a typical morphology of the NaCl crystal. It is also evident from the EDX analysis with higher counts of Na and Cl ions [see Fig. S4(III)]. The increment of the salts' presence at 12.5% (v/v), compared to 75% (v/v) is also evident from the micro-structural EDX analysis of the dried films [Figs. S4(III) and S5(III)]. Furthermore, the higher counts of Na and Cl than other ions in EDX micrographs confirm the excess concentration of NaCl in the adulterant (1x PBS).

While the structural deformation of these components (RBCs, WBCs, and platelets) provides a local response to the stress, the global response is achieved by the cracking of the blood droplet. The presence of the WBCs, the platelets, and some of the residual RBCs make the central region at 75% (v/v) crowded [see Figs. S4(I-II)]. The random cracks appear in this central region due to the lack of any dominant stress. As we add adulterant in the blood sample, the random cracks are replaced with the dendrite structure at 75% (v/v). The ring-like cracks are observed at 25% (v/v), whereas the $\phi$ of 12.5% (v/v) does not show any cracks in the central region under $5\times$ objective lens (Fig. **??**). This suggests that the various amount of salts creates different physio-chemical environments to release the mechanical stress in the central region. Due to high accumulation of salts, the clumping of cellular components are also found [see Fig. S5(I-II)]. The number of cracks increases in the salt-added diluted blood than the whole blood. However, this does not indicate that more stress has developed. It is just the fact that the energy cost of forming multiple shorter cracks (due to small corona width) is less than that of the single long crack (due to large corona width at $\phi = 100\%$). Interestingly, the sliding of the crack domains is only observed in the absence of the salts (bottom panel of Fig. S1 in the supplementary section). This suggests that the elastic network formed by the salts and the reduction of the blood's components result in the attachment of the dried droplet with the substrate.

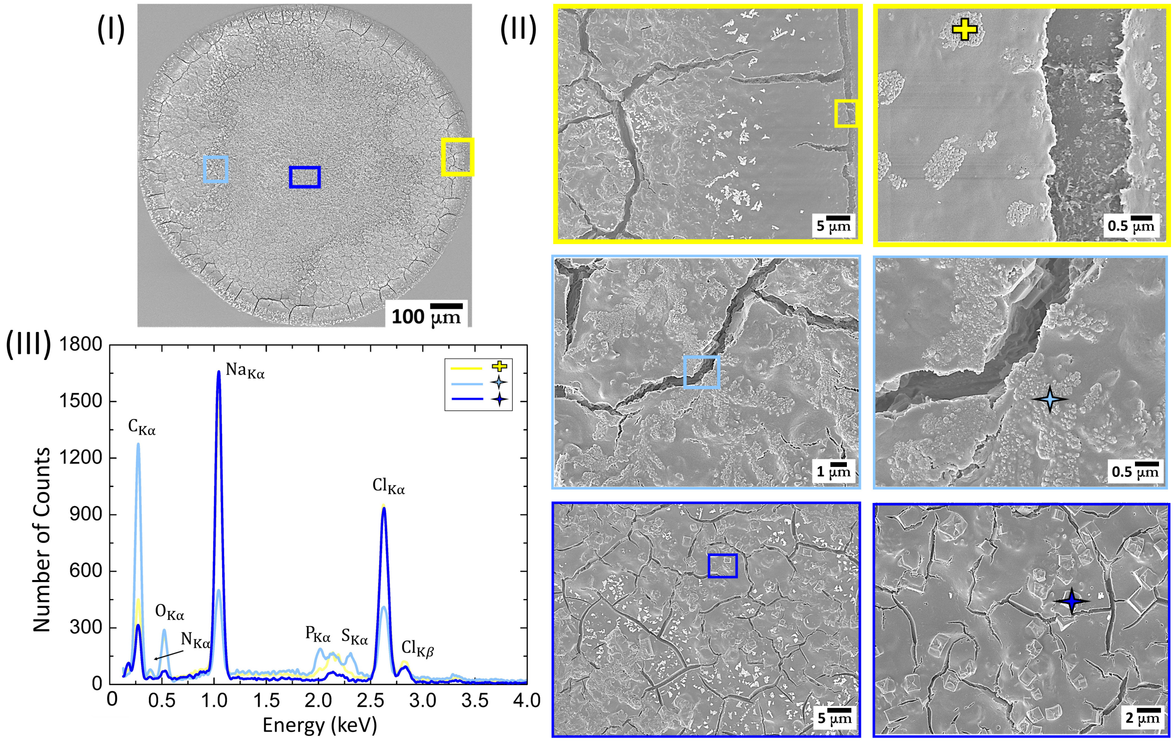


FIG. S5. The SEM images illustrating the microscopic structures at different length scales are shown in (I) and (II) at 75% (v/v). The yellow-colored rectangle in the top panel of (II) details the structures at the periphery and the corona. The light and dark blue colored rectangles in the middle and the bottom panels of (II) show different structures in the central regions. An EDX analysis shows the elemental composition of different regions of the blood-dried droplet in (III). The signals are symbolized and colored to match the area from where these were collected.

#### B. Machine learning (ML) for classifying the drying blood droplets

##### *1. ML to differentiate healthy, added water and added PBS blood droplets*

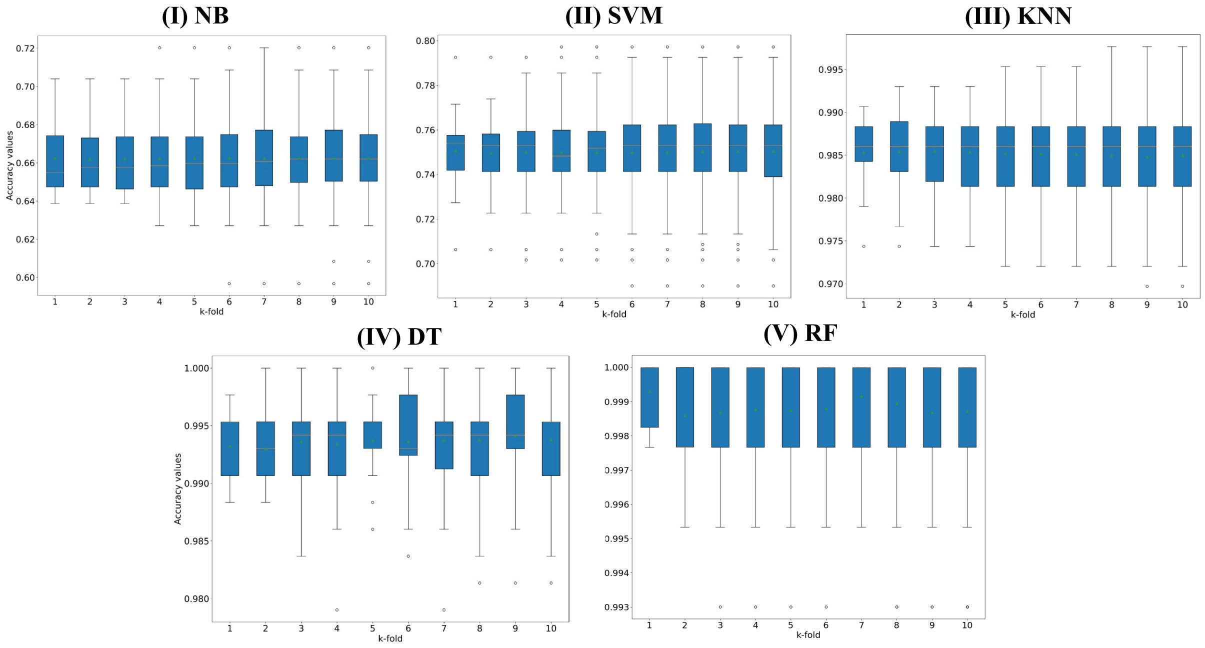


FIG. S6. Bar plots for the 10-fold cross validation as a function of number of folds (k = 10): (I) Naßve Bayes (NB), (II) Support Vector Machine (SVM), (III) K-Nearest Neighbors (KNN), (IV) Decision Tree (DT), and (V) Random Forest (RF).

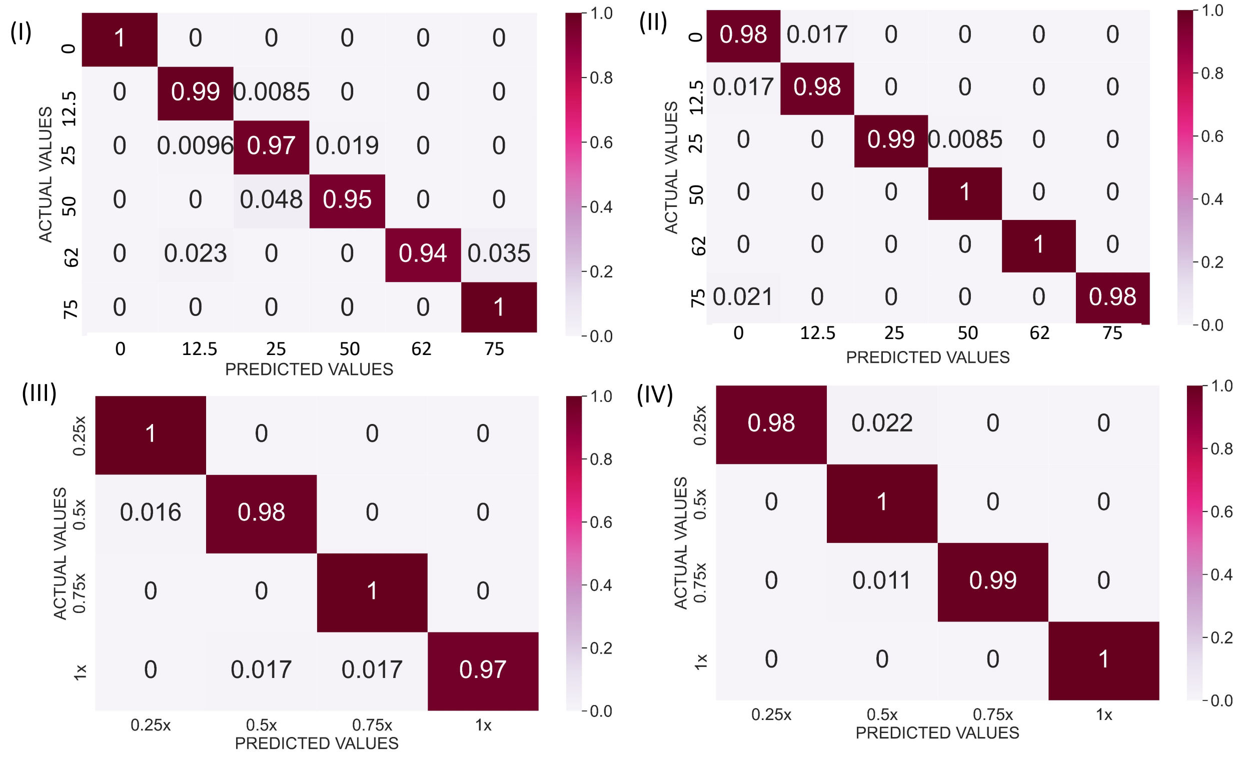


FIG. S7. Implementation of random Forest (RF) as MLA for classifying different adultered concentrations: Normalized confusion matrix for (I) blood+DI, (II) lys+PBS+LC, and (III) lys+PBS. The ROC curve for (IV) blood+DI, (V) lys+PBS+LC, and (VI) lys+PBS.

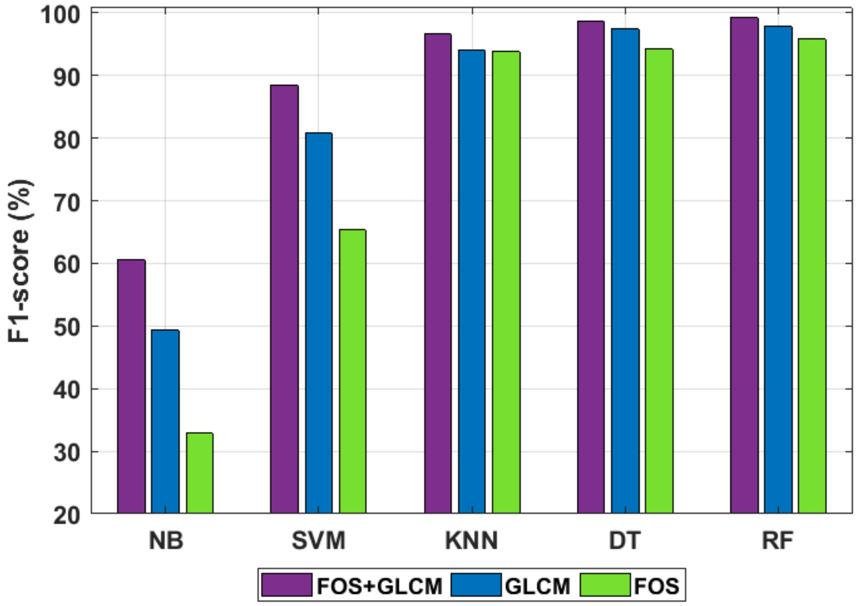


FIG. S8. Bar plots showcasing the class-wise performance of different traditional machine learning: Decision Tree (DT), Random Forest (RF), Support Vector Machine (SVM), K-Nearest Neighbors (KNN), and Naive Bayes (NB)..

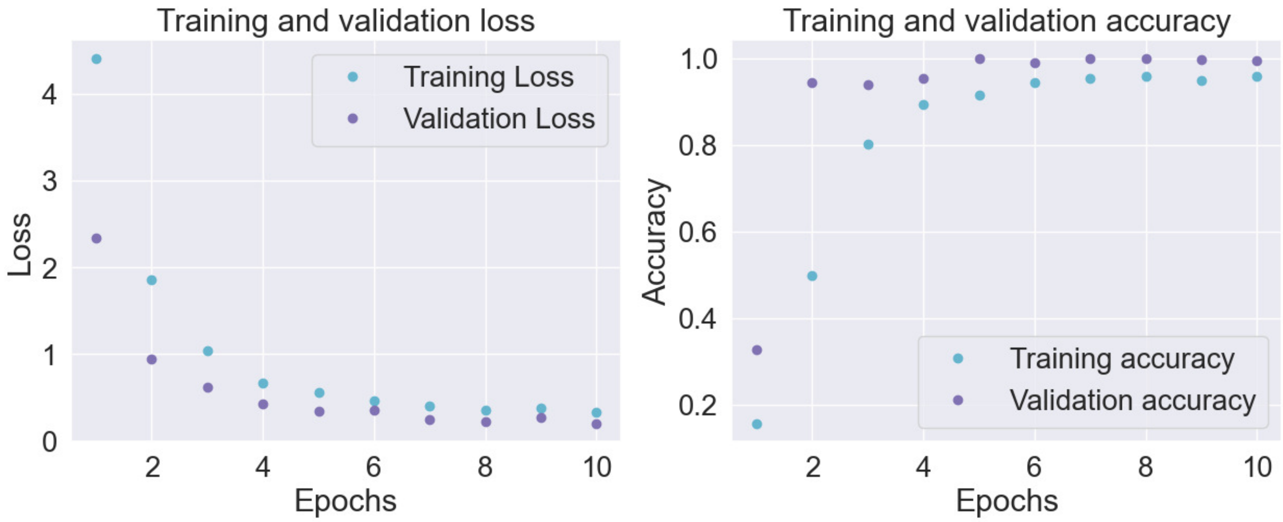


FIG. S9. Plots of the convolution neural network (CNN) using training and validation dataset: loss and accuracy of the model validated using a testing dataset and 11 classes.

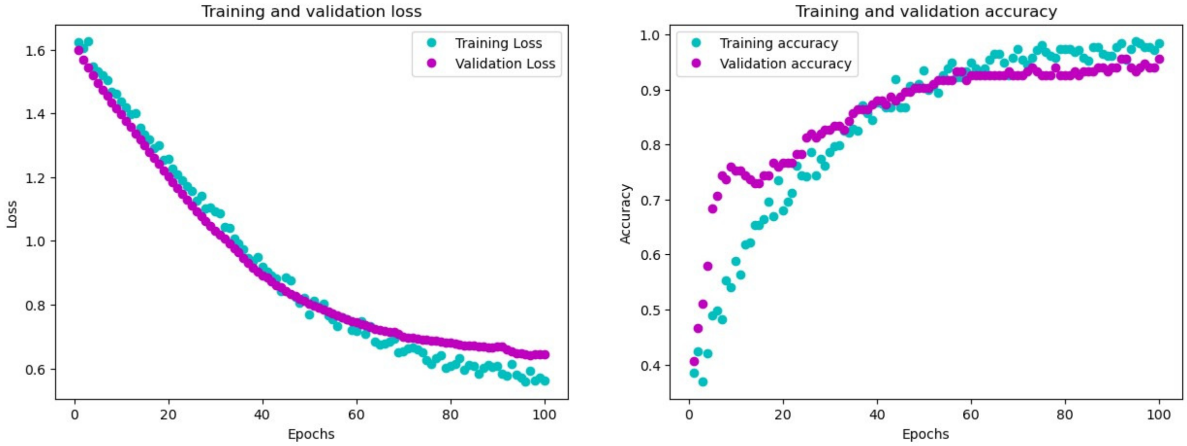


FIG. S10. Plots of the artificial neural network (ANN) using training and validation dataset: loss and accuracy of the model validated using a testing dataset and 11 classes.

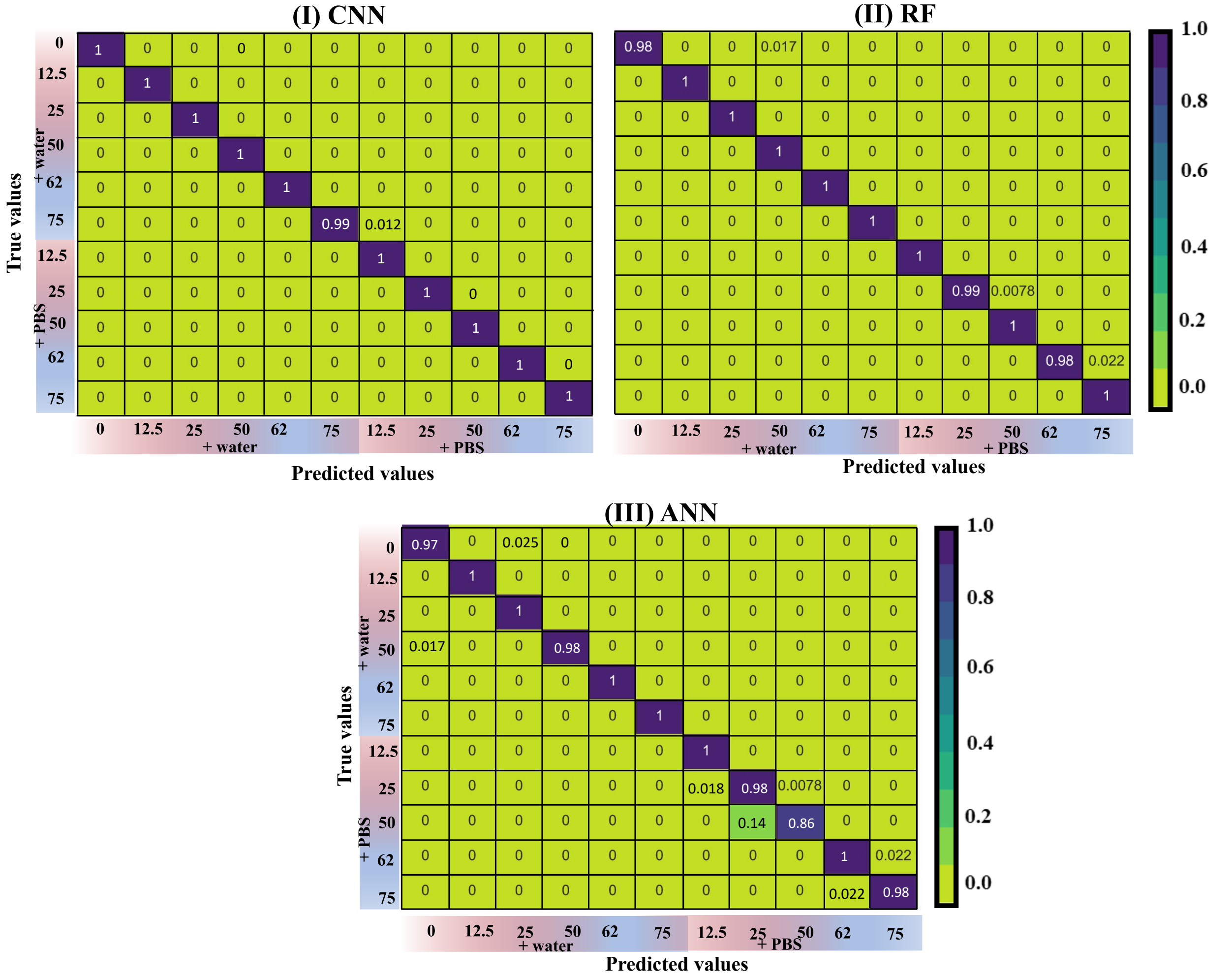


FIG. S11. Normalized confusion matrix of the (I) convolution neural network (CNN), (II) random forest (RF), and (III) artificial neural network (ANN) using a testing dataset to classify 11 blood samples, i.e., 0 (healthy), + water (12.5, 25. 50, 62, and 75 (v/v)%, and + PBS (12.5, 25. 50, 62, and 75 (v/v)%.

TABLE S1. Performance of Random forest as MLA on different bio-colloidal systems in the context of accuracy, precision, recall, and F1-score.

| (I) Blood+PBS | | | | (II) Blood+DI | | | |
|---|---|---|---|---|---|---|---|
| Concentrations | Precision | Recall | F1-score | Concentrations | Precision | Recall | F1-score |
| 100 (v/v) % | 1.000000 | 1.000000 | 1.000000 | 100 (v/v) % | 0.966942 | 0.983193 | 0.975000 |
| 75 (v/v) % | 0.975000 | 0.991525 | 0.983193 | 75 (v/v) % | 0.983051 | 0.983051 | 0.983051 |
| 62 (v/v) % | 0.943925 | 0.971154 | 0.957346 | 62 (v/v) % | 1.000000 | 0.991453 | 0.995708 |
| 50 (v/v) % | 0.980198 | 0.951923 | 0.965854 | 50 (v/v) % | 0.991150 | 1.000000 | 0.995556 |
| 25 (v/v) % | 1.000000 | 0.941860 | 0.970060 | 25 (v/v) % | 1.000000 | 1.000000 | 1.000000 |
| 12.5 (v/v) % | 0.974576 | 1.000000 | 0.987124 | 12.5 (v/v) % | 1.000000 | 0.979381 | 0.989583 |
| Accuracy | | | 0.978193 | Accuracy | | | 0.989247 |
| Macro average | 0.978950 | 0.976077 | 0.977263 | Macro average | 0.990191 | 0.989513 | 0.989816 |
| Weighted average | 0.978559 | 0.978193 | 0.978153 | Weighted average | 0.989362 | 0.989247 | 0.989270 |
| (III) Lys+PBS+LC | | | | (IV) Lys+PBS | | | |
| 0.25x | 0.983193 | 1.000000 | 0.991525 | 0.25x | 1.000000 | 0.977778 | 0.988764 |
| 0.50x | 0.983871 | 0.983871 | 0.983871 | 0.50x | 0.980000 | 1.000000 | 0.989899 |
| 0.75x | 0.981818 | 1.000000 | 0.990826 | 0.75x | 1.000000 | 0.989474 | 0.994709 |
| 1.00x | 1.000000 | 0.966102 | 0.982759 | 1.00x | 1.000000 | 1.000000 | 1.000000 |
| Accuracy | | | 0.987152 | Accuracy | | | 0.994398 |
| Macro average | 0.987221 | 0.987493 | 0.987245 | Macro average | 0.995000 | 0.991813 | 0.993343 |
| Weighted average | 0.987302 | 0.987152 | 0.987116 | Weighted average | 0.994510 | 0.994398 | 0.994403 |
